# Prematurity and Genetic Liability for Autism Spectrum Disorder

**DOI:** 10.1101/2024.11.20.24317613

**Authors:** Yali Zhang, Ashraf Yahia, Sven Sandin, Ulrika Åden, Kristiina Tammimies

**Affiliations:** Center of Neurodevelopmental Disorders (KIND), Centre for Psychiatry Research, Department of Women’s and Children’s Health, Karolinska Institutet; Astrid Lindgren Children’s Hospital, Karolinska University Hospital, Region Stockholm, Stockholm, Sweden; Department of Medical Epidemiology and Biostatistics, Karolinska Institutet, Stockholm, Sweden; Department of Psychiatry, Icahn School of Medicine at Mount Sinai, New York, USA; Seaver Center for Research and Treatment at Mount Sinai, New York, USA; Department of Women’s and Children’s Health, Karolinska Institutet, Stockholm, Sweden; Department of Neonatology, Division of Neonatal Medicine, Karolinska University Hospital, Stockholm, Sweden; Department of Bioclinical sciences, Linköping University, Linköping, Sweden

**Keywords:** Prematurity, Autism Spectrum Disorder, Genetics, Polygenic risk score, Machine learning, Generalized estimating equations model

## Abstract

**Background:** Autism Spectrum Disorder (ASD) is a neurodevelopmental condition characterized by diverse presentations and a strong genetic component. Environmental factors, such as prematurity, have also been linked to increased liability for ASD, though the interaction between genetic predisposition and prematurity remains unclear. This study aims to investigate the impact of genetic liability and preterm birth on ASD conditions.

**Methods:** We analyzed phenotype and genetic data from two large ASD cohorts, the Simons Foundation Powering Autism Research for Knowledge (SPARK) and Simons Simplex Collection (SSC), encompassing 78,559 individuals for phenotype analysis, 12,519 individuals with genome sequencing data, and 8,104 individuals with exome sequencing data. Statistical significance of differences in clinical measures was evaluated between individuals with different ASD and preterm status. We assessed the rare variants burden using generalized estimating equations (GEE) models and polygenic load using ASD-associated polygenic risk score (PRS). Furthermore, we developed a machine learning model to predict ASD in preterm children using phenotype and genetic features available at birth.

**Results:** Individuals with both preterm birth and ASD exhibit more severe phenotypic outcomes despite similar levels of genetic liability for ASD across the term and preterm groups. Notably, preterm ASD individuals showed an elevated rate of de novo variants identified in exome sequencing (GEE model, *p*=0.005) in comparison to the non-ASD preterm group. Additionally, a GEE model showed that a higher ASD PRS, preterm birth, and male sex were positively associated with a higher predicted probability for ASD, reaching a probability close to 90% in SPARK. Lastly, we developed a machine learning model using phenotype and genetic features available at birth with limited predictive power (AUROC = 0.65).

**Conclusions:** Preterm birth may exacerbate the multimorbidity present in ASD, which was not due to the ASD genetic factors. However, increased genetic factors may elevate the likelihood of a preterm child being diagnosed with ASD. Additionally, a polygenic load of ASD-associated variants had an additive role with preterm birth in the predicted probability for ASD, especially for boys. We propose that incorporating genetic assessment into neonatal care could benefit early ASD identification and intervention for preterm infants.

## Introduction

Autism Spectrum Disorder (ASD) is an early-onset neurodevelopmental condition characterized by challenges in social interaction, communication, and restrictive and repetitive behaviors and interests [1]. In addition to these core symptoms, individuals with ASD have multiple co-occurring neurodevelopmental, psychiatric, and physical conditions, which contribute to clinical heterogeneity [2].

The etiology of ASD is multifaceted and not yet fully elucidated [3,4]. However, genetic factors account for up to 80-90% of the liability for ASD [4–6]. Rare de novo variants (DNV), especially those affecting the gene function of constraint genes, are shown to be enriched in ASD [7,8]. Rare inherited variants in ASD-related genes are also shown to be overtransmitted from parents to their children with ASD [5,9]. In addition to rare variants, genome-wide association studies (GWAS) have identified a few common variants associated with ASD, and the polygenic load calculated using polygenic risk score (PRS) has demonstrated predictive ability for ASD and ASD traits [10,11], explaining 2% of variance in ASD status [12]. Furthermore, ASD PRS can uniquely predict variability in cognitive performance [11].

In addition to genetic factors, there are several environmental factors associated with ASD [3]. The most robustly associated environmental stressor is prematurity, with ASD likelihood in preterm about two to four folds higher than in term, and ASD likelihood increasing as gestational age at birth decreases [13,14]. Although preterm birth involves both genetic and environmental components [15], it is typically discussed as an environmental factor in ASD studies [3,16]. Preterm birth, defined as delivery with gestational age before 37 weeks, can be further categorized into four preterm sub-categories: extremely preterm (<28 weeks), very preterm (28-31 weeks), moderate preterm (32-33 weeks), and late preterm (34-36 weeks). Prematurity is not only associated with ASD but also with neurocognitive development and other health outcomes [17–19]. Earlier studies investigating phenotypes in children with ASD suggested that extremely or very preterm ASD children have more language deficits and developmental delays compared to term ASD children [20,21]. However, others reported that no significant differences in development were found when studying the entire preterm group [22]. Moreover, preterm birth as an exposure is associated with various comorbidities in ASD, including attention and behavioral problems, neurological disorders, and growth deficiency. However, more investigations are needed to understand ASD phenotypic spectrum in preterm and term birth as well as how different sub-groups of prematurity contribute to specific medical outcomes and traits.

Interestingly, preterm infants have been found to have increased DNV rates compared to term [23], and various de novo copy number variations (CNVs) in preterm were found related to neurodevelopmental disorders genes (NDD genes) [24], but it remains uncertain whether DNV burden is further elevated when both ASD and preterm birth are present. Moreover, the relationship between ASD polygenic load and prematurity has only been evaluated by Cullen et al based on cognition, but no interaction was found between ASD PRS and gestational age at birth [16]. Furthermore, there are indications that a small fraction of preterm individuals would have recognizable genetic disorders [25]. However, there have not been specific studies focusing on genetic factors within preterm individuals and ASD.

While genetic and phenotypic studies on the population level are informative, these analyses may miss interactions and non-linear relationships within or between factors on an individual level. To address this complexity, machine learning (ML) has the potential to identify patterns in high-dimensional data that traditional statistical methods may overlook, aiding in prediction. To date, ML prediction models have emerged to predict ASD using different data sources, such as routine medical assessments and electronic records [26,27], genetic data [28], and integrative models [29]. However, none of the published ML models currently predict ASD in preterm children. In the existing ML models for ASD prediction, included features are typically collected when the child is at least 1-2 years of age or older [26,30]. It remains unclear whether integrating phenotype and genetic information available at birth could enable earlier ASD identification in preterm infants.

In this study, we aimed to enhance our understanding of ASD in preterm children by analyzing both clinical and genetic data in two large ASD cohorts, the Simons Foundation Powering Autism Research for Knowledge (SPARK) [8,31], and the Simons Simplex Collection (SSC) [32]. Across individuals with different ASD and prematurity sub-groups, we first examined their phenotype severity through the prevalence and multimorbidity of other medical diagnoses. Thereafter, we assessed the burden of rare and common sequence-level variants. Finally, we built an ML model using both phenotype and genetic features that could be obtained at birth to predict ASD in preterm individuals.

## Methods

### Study cohorts

The Simons Foundation Powering Autism Research for Knowledge (SPARK) database, initiated by the Simons Foundation Autism Research Initiative (SFARI), recruited families in the USA with one or more children diagnosed with autism spectrum disorder (ASD) [31]. We utilized demographic and phenotype data from the SPARK collection version 9 with a release date 2022-12-12. We considered medical and psychiatric diagnosis history from the basic medical screen dataset, grouping specific diagnoses into nine diagnostic categories: behavior, development, mood, growth, birth, eating habits (Eat), neurological conditions (Neuro), visual and auditory impairments (Visaud), and sleep. This dataset includes 9,196 individuals with ASD and born preterm with gestational age less than 36 weeks (ASD-preterm), 65,021 individuals with ASD and born term (ASD-term), and 2,706 individuals without ASD and born preterm (non-ASD-preterm). We also stratified preterm individuals into four sub-groups based on gestational age: extremely preterm (<28 weeks), very preterm (28-31 weeks), moderate preterm (32-33 weeks), and late preterm (34-36 weeks). Additionally, we compared quantitative measures using the Child Behavior Checklist (CBCL) t-score for 1 to 5 and 6 to 18 years of age, Developmental Coordination Disorder Questionnaire (DCDQ) final score, Repetitive Behavior Scale-Revised (RBS-R) total final score, Social Communication Questionnaire (SCQ) final score and Full-Scale Intelligence quotient (FSIQ) score. Additional file 1: Tables S1 and S2 provide detailed description of the specific diagnoses and quantitative measures.

For the genetic part, dataset SPARK genome sequencing (GS) version 1.1 variant calling was used, including 12,519 individuals from 3,394 families, with 315 ASD-preterm, 2,788 ASD-term, and 155 non-ASD-preterm individuals. We also utilized earlier published DNV data from exome sequencing (ES) to calculate the event rate [8]. Among the 6,444 ASD individuals with DNV information, 5,747 were born full-term, and 697 were born preterm. Furthermore, DNV information was accessible for 210 preterm children without ASD.

We also used the Simons Simplex Collection (SSC) cohort. SSC recruited more than 10,000 individuals from 2,000 families [32]. Due to the absence of preterm information for non-ASD individuals (siblings of ASD probands), we only conducted studies for ASD-preterm and ASD-term groups. After excluding individuals with unreliable gestational age, unknown ASD diagnosis, births occurring post-term (gestational age > 40 weeks), and missing outcomes information, we retained 1,637 probands diagnosed with ASD (157 preterm and 1,479 term) for the diagnostic category analysis. Additionally, we analyzed available quantitative measures, including CBCL score, DCDQ score, SCQ score, and IQ score. Detailed descriptions of specific diagnoses and qualitative measures are provided in Additional file 1: Tables S3 and S4. We incorporated the de novo variants (DNV) dataset from Ng et al in our analysis, encompassing 1,450 ASD individuals after excluding post-term births [33]. Additionally, the Polygenic Risk Score (PRS) dataset we used sourced from Weiner et al comprised 1,590 ASD individuals, excluding post-term births [34].

### De novo variant calling and analysis

In SPARK, GS was conducted on the Illumina NovaSeq 6000 system. Variant calling was performed using GATK (v3.5) with HaplotypeCaller, and all samples were jointly called by GLnexus (v1.4.1). To find de novo variants (DNV) of children, we included all trios. For families with more than one child, each child forms a trio with their parents, resulting in multiple trios within the same family. There were 5,712 trios from 3,364 families. We used two tools, Slivar (v0.2.8) and GATK (v4.1.4.1), to call the DNV from SPARK trios, and true DNV was selected when it was found in both tools [35,36]. The DNV was called if it was labeled as “denovo” with allele balance (AB) in children higher than 0.25 in Slivar and identified as high confidence DNV in GATK. For pseudo-autosomal regions on the sex chromosome, we separately considered the variant genotype as 1/0 in children. We did not find DNVs on chrY in pseudo-autosomal regions. Then, we did quality control to further filter the rare DNV by removing variants with GQ<20, DP<10, gnomAD population allele frequencies > 0.001, and variants of either 10 A’s or T’s in a row. We filtered out DNVs on genomic centromeres and low complex regions. Then we removed DNV if 1) it can be found in other family’s parents, 2) it can be found in only children but more than three families, and 3) it on positions having more than 3 multi-alleles. We identified 432,903 DNVs, including 16,155 exonic DNVs and 986 loss-of-function (LOF) variants. We filtered out 29 children with DNV counts beyond three times the standard deviation from the mean DNV count. Based on these criteria, the average number of rare DNVs per child was 75.9.

We annotated DNVs by ANNOVAR (v2020.06.08) with buildver as hg38 and protocol as ljb26_all, SnpEff (v5.2) with GRCh38.105 databases, and VEP (v110) with assembly as GRCh38 and 154 dbSNP [37–39]. DNVs with Sequence Ontology (SO) terms as "frameshift", "splice_acceptor", "splice_donor", "start_lost", "stop_gained", and "stop_lost" in gene effect were considered as LOF DNV. Additionally, we identified variants on the neurodevelopmental disorder-related genes (NDD gene) using high-confidence ASD genes collected by the Simons Foundation Autism Research Initiative (SFARI) (2024-01-16 release) with gene scores of 1 or 2 and labeled as syndromic and green gene list of Intellectual disability - microarray and sequencing (Version 5.497) on Genomics England PanelApp (2024-03-14 accessed) (Additional file 2) [40,41].

### Inherited variants calling

Using the same GS data as for DNV calling, including 5,712 trios from 3,364 families, we extracted variants on the genomic protein-coding NDD genes (Additional file 2) using VCFtools (v0.1.16) [42]. Then we annotated the inheritance mode of NDD genes using the ID gene panel app, SysNDD database (v0.1.0) and DDgenes [41,43,44]. There were 80% (1625 genes) of NDD genes annotated, including 738 dominant genes coded as monoallelic or dominant, and 944 recessive genes coded as biallelic or recessive in databases (Additional file 2). To restrict the analysis to rare inherited variants, we used the allele frequency filter threshold of 0.001 and 0.01 for dominant and recessive genes, respectively. Variants with GQ<20, DP<10, and genotypes conflicting with the inheritance mode of located genes were filtered out. We did not find compound variants (more than 1 heterozygous variant on the same recessive gene for one child). For variants on dominant genes, we identified LOF following the same process in the DNV part and found damaging missense variants met at least one of the following conditions: CADD>=20, SIFT labeled as D, POLYPHEN labeled as P and D, PHYLOP>=2.0 or REVEL>=0.5. From 5,712 trios in the SPARK GS database, we identified 245,671 and 4,346 inherited variants in dominant and recessive NDD genes, respectively. We identified 2,717 LOF and 39,136 damaging missense variants in genes with a dominant inheritance mode.

### Polygenic risk score

Based on GS jointly called variants from 12,519 SPARK participants, we performed quality control for individuals and variants using PLINK1.9 with parameters listed in Additional file 1: Table S5, retaining 11,933 individuals and 9,558,997 variants with a total genotyping rate of 99.9% [45]. Then, genome coordinates of variants were converted from hg38 to hg19 using liftOver (v2017-03-14) [46], and 9,428,216 were mapped after removing duplicated variants. We calculated the posterior SNP effect size estimates using PRS-CS with ASD GWAS summary statistics from the Psychiatric Genomics Consortium (November 2017 release) (46,351 individuals) and European LD reference data from 1000 Genome phase III [10,47]. The default parameters used in PRS-CS also be listed in Additional file 1: Table S5. The final PRS was calculated using the score function in PLINK1.9 with the estimated posterior SNP effect size. To minimize the effects on different populations, we analyzed the ancestry of individuals using Principal component analysis (PCA) by pca command in PLINK1.9. The ten principal components (PC1-10) were included as covariates when we calculated the association between phenotype and PRS.

### Statistical analysis

All analyses were performed using R programming language (v4.2.2). Commencing with an exploration of phenotypes, we investigated two pairs of groups: preterm and term birth within individuals diagnosed with ASD (ASD-preterm versus ASD-term); and individuals diagnosed with ASD to those without ASD within preterm birth (ASD-preterm versus non-ASD-preterm). An individual was considered to possess the diagnosis feature if any specific diagnosis within that diagnostic category was exhibited, no matter how many specific diagnoses there were at the same time.

In phenotypic analysis, the prevalence is reported by the frequency of individuals with the diagnostic categories (Additional file 1: Table S1). We examined the differences in prevalence between ASD-preterm and ASD-term, ASD-preterm and non-ASD-preterm using odds ratios (ORs) with 95% confidence interval (CI) and statistical significance reported by FDR-adjusted p-values. After stratifying preterm stages, we used χ² test to evaluate differences across preterm stages, post-hoc comparisons for each pair of preterm stages, and Kendall’s tau test to examine rank correlation of preterm stages with prevalences. Then, prevalences and ORs of multimorbidity (one, two, three, four, or not less than five diagnostic categories; Additional file 1: Tables S1) were estimated by ASD and preterm status, and prevalence differences in multigroup were examined by χ² test. We also examined prevalence and multimorbidity ORs adjusted for sex and age using generalized estimating equation (GEE) models, with family ID as a clustering variable. GEE model is more robust to assumptions of data following a particular data distribution and adjusts for correlations between individuals, e.g., siblings and families [48]. Additionally, we examined differences in quantitative measures using the 2-sided Wilcoxon rank sum test for pairwise comparisons and in multiple comparisons using the Kruskal-Wallis rank sum test. To account for multiple testing, we applied false discovery rate correction to p-values.

The burden of DNV and inherited variants was evaluated by comparing rates of such variants in each subgroup categorized by ASD and preterm status. We assessed the statistical differences between groups through GEE models with Poisson family, sex as a covariate, and family ID as a clustering variable. PRSs were z-standardized and statistical significance for PRS distribution was reported by the 2-sided Wilcoxon rank sum test. The association between targeted phenotype (y/n) and PRS in each subgroup was also evaluated in GEE logistic model with sex(m/f) and PC1-10 from ancestry checking as covariates.

The variance of ASD status explained by PRS in the SPARK cohort was quantified using the McFadden Pseudo-r^2^ value based on the logistic regression model adjusted for sex and ancestry PC1-10. To examine the associations between ASD diagnosis and possible variables, we modeled the probability of ASD (y/n) by fitting GEE logistic model(s) with the equation as [ASD (y/n) ∼ sex (m/f) + preterm (y/n) + standardized PRS] and family ID as a clustering variable in European population to find the correlation between ASD diagnosis and possible variables. To visualize the predicted probabilities of ASD from the GEE logistic model, we utilized ggemmeans function in ggeffects R package (v1.5.1) [49], showing the average predicted probabilities of ASD for specific levels of variables adjusted for other covariates in the model. After that, the variable (preterm (y/n) * standardized PRS) was added to the GEE logistic model to check the correlation between ASD (y/n) and the interaction of preterm status and PRS. To detect the association between multimorbidity and DNV burden, GEE models (Table S4) and linear regression were used.

### Machine learning model

This part of the analysis was performed in R (v4.2.2). Within preterm individuals, we utilized non-ASD and ASD diagnoses as two classification flags, incorporating features obtained from data that can be collected at birth. For phenotypical variables, we included birth complications, sex, and birth-related conditions. Genetic variables encompassed the count of several types of genetic variants, the CADD score of de novo variants (DNV), and the PRS of ASD. To remove redundant variables and identify informative features, we employed the Recursive Feature Elimination (RFE) algorithm [50]. RFE is a feature selection technique that iteratively removes the least important features based on model performance, refitting the model with the remaining features until the optimal subset of features is identified. In RFE, we utilized random forest function and 10-fold cross-validation in the underlying model to assess feature importance throughout the process. For the features selected after RFE, we only retained the more general feature (e.g., retaining LOF over LOF on NDD genes) in any pair of features with a correlation coefficient above 0.7 to reduce multicollinearity. The details of selected features are listed in Additional file 1: Table S6.

We applied R package caret (v6.0-94) [51] to train the ML models. Given the limited sample size and the higher proportion of ASD samples than non-ASD, we conducted nested cross-validation (NCV) and hyperparameter tuning with grid search to enhance model performance. In NCV, we partitioned the data into 10 folds in the outer loop, with nine folds used for training and the remaining fold for testing. Within the inner loop, we performed repeated 10*10-fold cross-validation, and the model with the best performance was applied to the outer loop. We employed three algorithms—Extreme Gradient Boosting (XGBoost), Random Forest (RF), and Linear Support Vector Machine (SVM)—to construct the models. The values of hyperparameter tuning for each model are detailed in Additional file 1: Table S7. We reported evaluation metrics, including accuracy with a 95% CI, area under the receiver operating characteristic curve (AUROC), specificity, sensitivity, and F1-score. Moreover, the SHapley Additive exPlanations (SHAP) values for features were computed and visualized using the R package SHAPforxgboost (v0.1.3) [52], quantifying the contribution of each feature to individual model predictions in terms of direction and magnitude.

## Result

### Phenotype comparison across ASD and prematurity

We utilized basic medical screening data from 181,248 individuals in the SPARK version 9 cohort (release date 2022-12-12). Among them, 74,217 (41%) were diagnosed with ASD, and 11,902 (7%) were born preterm. When performing phenotype comparison, we grouped individuals with 9,196 individuals with ASD and being preterm (ASD-preterm), 65,021 individuals with ASD but being term (ASD-term), and 2,706 preterm individuals without ASD diagnosis (non-ASD-preterm) (Figure 1, Table 1). In the SSC cohort, gestational age records were available only for probands with ASD, of which 157 were preterm and 1,479 were term. We stratified the preterm stage based on gestational age at birth (Table 1).

**Figure 1.**
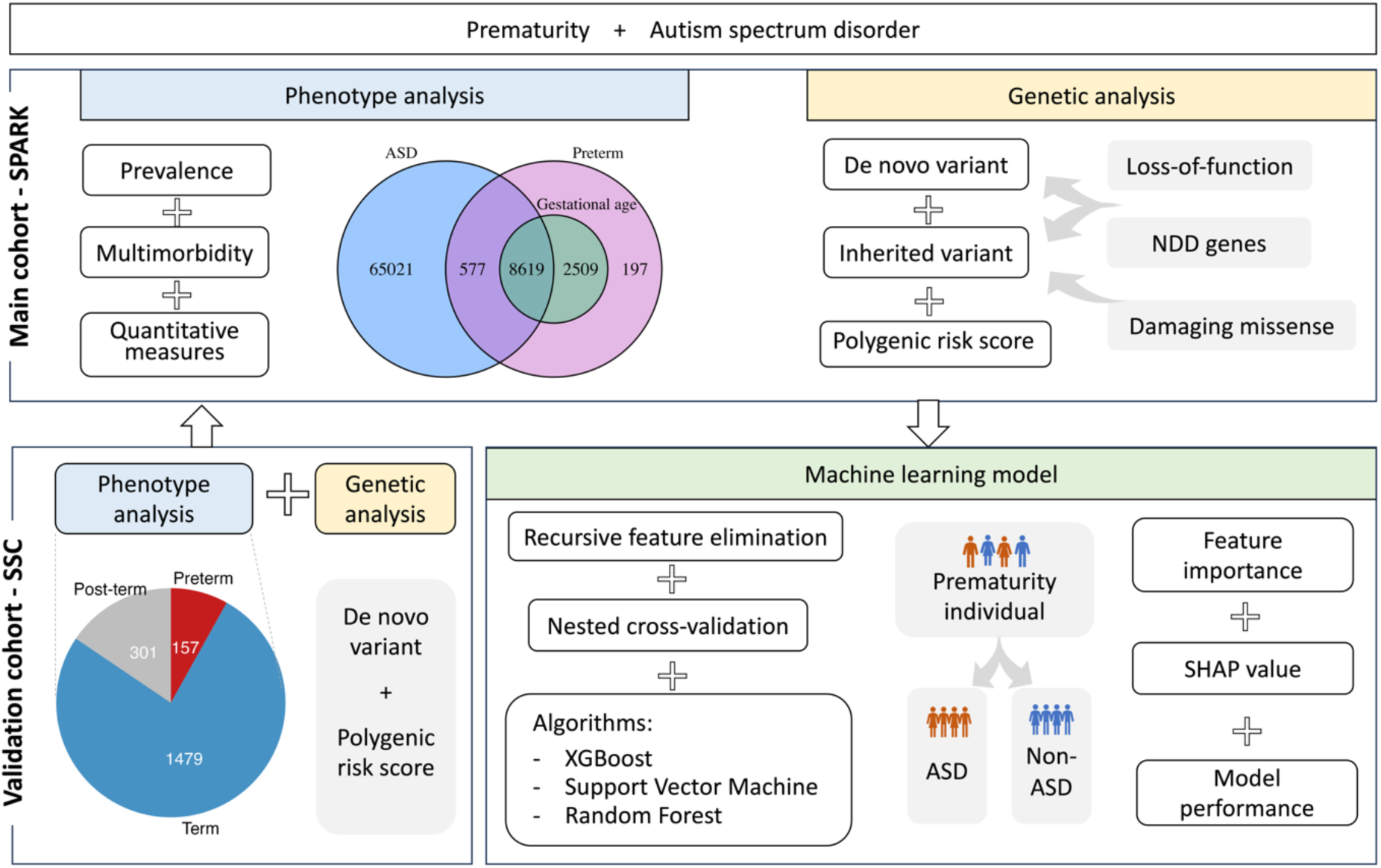
Overview design of the study. Firstly, we performed phenotype analysis on diagnosis prevalence, burden of multimorbidity and quantitative measures in the SPARK cohort. The sample size of SPARK is shown in the Venn diagram, with blue indicating ASD, pink indicating preterm with unknown gestational age, and green indicating preterm with known gestational age. Secondly, we analyzed the de novo variant and inherited variant burden, separately, focusing on loss-of-function variants and damaging missense and if these affected neurodevelopmental disorder (NDD) genes. Additionally, we utilized polygenic risk scores for common variants associated with ASD. For validation, we applied similar analyses in the SSC cohort. Thirdly, we integrated phenotype and genomic data to train the machine learning models with different algorithms to predict ASD diagnosis in the preterm group. Shapley additive explanations (SHAP) values assess the effect of each feature on the model performance.

**Table 1.**
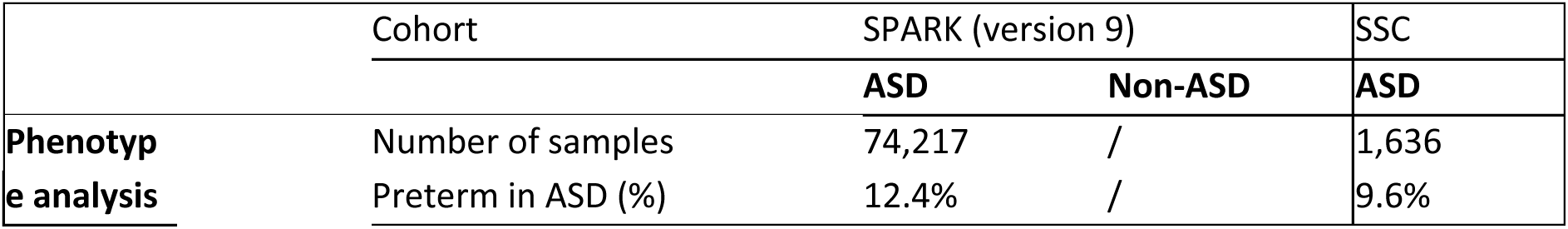
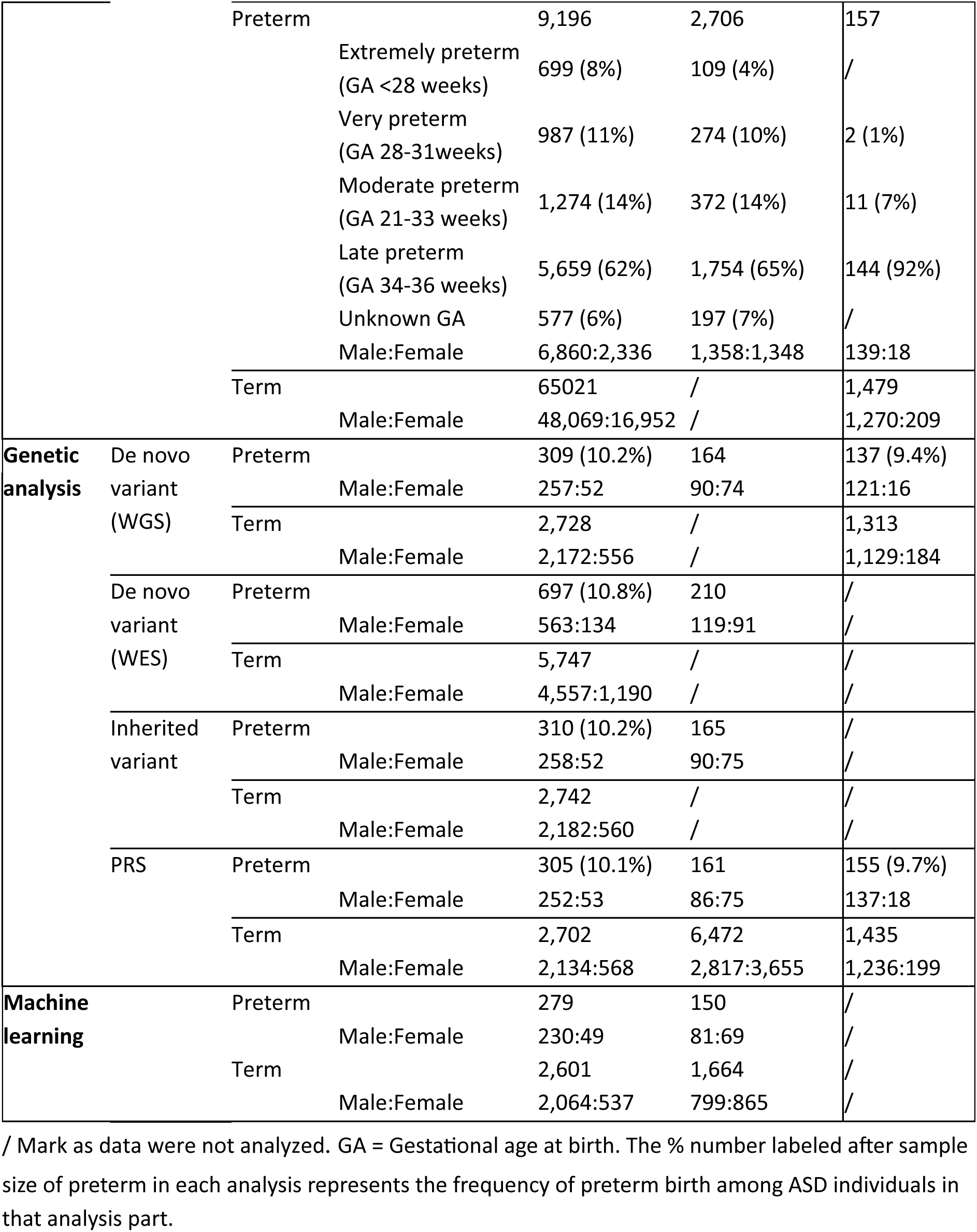
Characteristics of the analyzed participants.

|  | Cohort | SPARK (version 9) |  | SSC |
| --- | --- | --- | --- | --- |
|  |  | ASD | Non-ASD | ASD |
| **Phenotypic analysis** | Number of samples | 74,217 | / | 1,636 |
|  | Preterm in ASD (%) | 12.4% | / | 9.6% |

|  |  |  |  |  |  |
| --- | --- | --- | --- | --- | --- |
|  |  | Preterm | 9,196 | 2,706 | 157 |
|  |  | Extremely preterm<br>(GA <28 weeks) | 699 (8%) | 109 (4%) | / |
|  |  | Very preterm<br>(GA 28-31weeks) | 987 (11%) | 274 (10%) | 2 (1%) |
|  |  | Moderate preterm<br>(GA 21-33 weeks) | 1,274 (14%) | 372 (14%) | 11 (7%) |
|  |  | Late preterm<br>(GA 34-36 weeks) | 5,659 (62%) | 1,754 (65%) | 144 (92%) |
|  |  | Unknown GA | 577 (6%) | 197 (7%) | / |
|  |  | Male:Female | 6,860:2,336 | 1,358:1,348 | 139:18 |
|  |  | Term | 65021 | / | 1,479 |
|  |  | Male:Female | 48,069:16,952 | / | 1,270:209 |
| **Genetic analysis** | De novo variant<br>(WGS) | Preterm | 309 (10.2%) | 164 | 137 (9.4%) |
|  |  | Male:Female | 257:52 | 90:74 | 121:16 |
|  |  | Term | 2,728 | / | 1,313 |
|  |  | Male:Female | 2,172:556 | / | 1,129:184 |
|  | De novo variant<br>(WES) | Preterm | 697 (10.8%) | 210 | / |
|  |  | Male:Female | 563:134 | 119:91 | / |
|  |  | Term | 5,747 | / | / |
|  |  | Male:Female | 4,557:1,190 | / | / |
|  | Inherited variant | Preterm | 310 (10.2%) | 165 | / |
|  |  | Male:Female | 258:52 | 90:75 | / |
|  |  | Term | 2,742 | / | / |
|  |  | Male:Female | 2,182:560 | / | / |
|  | PRS | Preterm | 305 (10.1%) | 161 | 155 (9.7%) |
|  |  | Male:Female | 252:53 | 86:75 | 137:18 |
|  |  | Term | 2,702 | 6,472 | 1,435 |
|  |  | Male:Female | 2,134:568 | 2,817:3,655 | 1,236:199 |
| **Machine learning** | Preterm |  | 279 | 150 | / |
|  |  | Male:Female | 230:49 | 81:69 | / |
|  | Term |  | 2,601 | 1,664 | / |
|  |  | Male:Female | 2,064:537 | 799:865 | / |

/ Mark as data were not analyzed. GA = Gestational age at birth. The % number labeled after sample size of preterm in each analysis represents the frequency of preterm birth among ASD individuals in that analysis part.

We performed analysis on nine available diagnostic categories recorded in the basic medical screening dataset in SPARK phenotype database version 9, involving behavior, development, mood, growth, birth, eating habits (eat), neurological conditions (neuro), visual and auditory impairments (visual), and sleep (details of diagnostic categories are described in Additional file 1: Table S1). For each category, we assigned a binary variable indicating the presence or absence of conditions within that category, rather than counting the number of specific diagnoses. The prevalence of all diagnostic categories analyzed was higher in ASD-preterm compared to ASD-term (Figure 2A). Specifically, ASD-preterm had higher odds ratio (OR) for all diagnostic categories, with the highest being for birth and growth diagnoses (ORs=2.18 [95% CI 1.97–2.40] and 2.18 [95% CI 2.06–2.31], respectively). Additionally, preterm birth was associated with a modestly increased likelihood of other behavioral diagnoses compared to term in the ASD group (OR=1.19 [95% CI 1.14–1.25]). The results remained consistent after adjusting for sex, age, and family linkage using GEE models (Additional file 1: Table S8). We also observed significantly different prevalences of diagnostic categories (χ² test with FDR-adjusted *p*<0.001 for all categories) when we considered different sub-groups of preterm birth (Additional file 1: Figure S1A). Furthermore, we identified linear trends across different preterm stages, with groups of lower gestational age showing a higher prevalence in growth, eating, neuro, and visual diagnostic categories (Kendall’s tau test, FDR-adjusted *p*=0.04). Almost all the preterm sub-groups had a higher prevalence of diagnostic categories compared to the term stage (FDR-adjusted *p* of the post-hoc comparisons of χ² test are in Additional file 1: Table S9)

**Figure 2.**
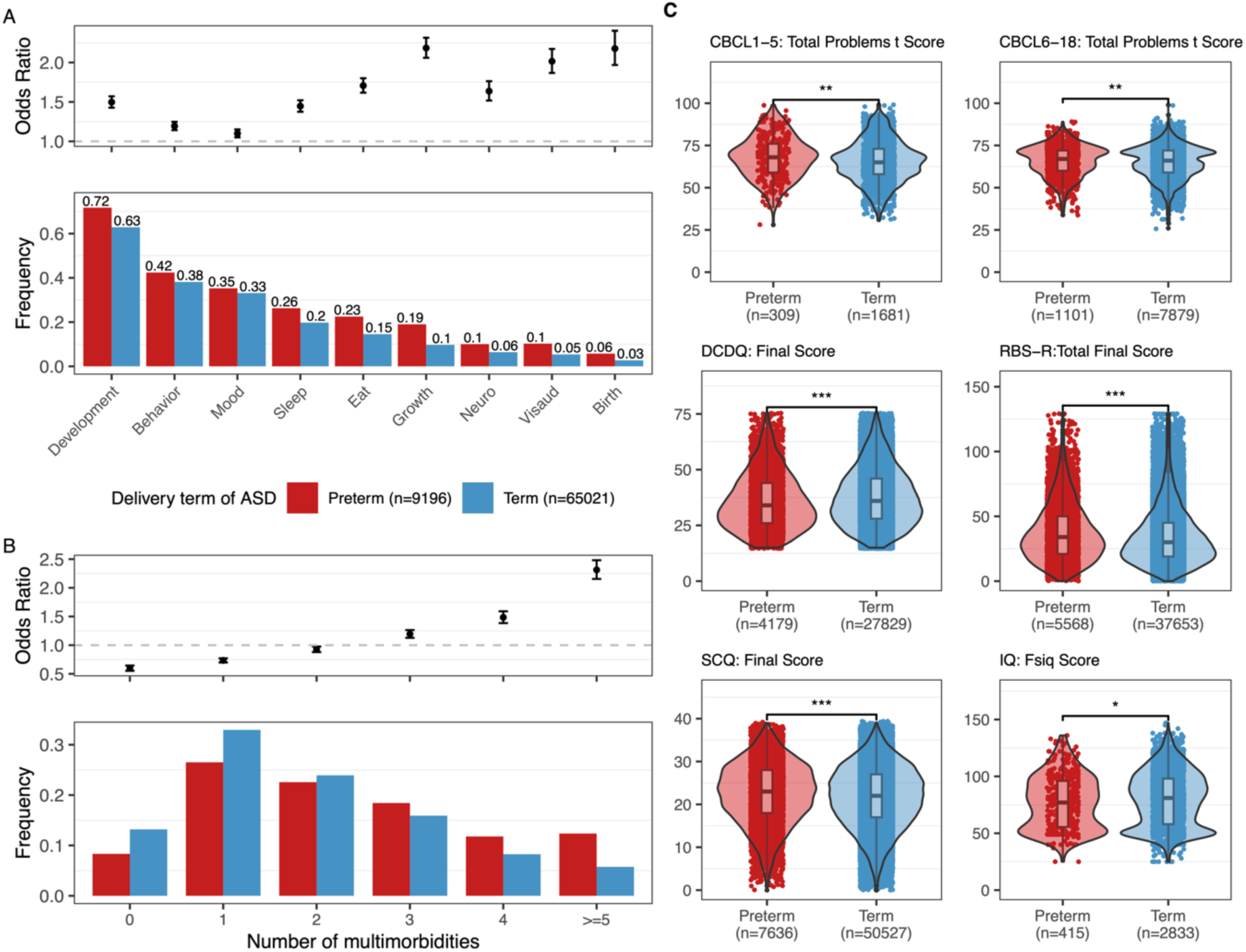
The phenotype comparison between preterm and term with ASD in the SPARK version 9 cohort. Color bars are the same across three panels and shown at the bottom of panel A. A. Prevalence and odds ratio with 95% confidence interval (CI) of diagnostic categories. The detailed outcomes in each diagnostic category are described in Table S1. The exact prevalence values are labeled on the top of the bars. ORs are given among ASD individuals born preterm vs term and recorded in Table S8. B. Distribution of the number of multimorbidities, defined as the total number of diagnostic categories that appear in individuals. ORs with 95% CI are given among ASD individuals born preterm vs term when focusing on each multimorbidity number and recorded in Table S10. C. Differences in Child Behavior Checklist (CBCL) t-score for 1 to 5 and 6 to 18 years of age, Developmental Coordination Disorder Questionnaire (DCDQ), Repetitive Behavior Scale-Revised (RBS-R) score, Social Communication Questionnaire (SCQ) and Full-Scale IQ (Fsiq) among ASD individuals born preterm and term. Significance was assessed using the 2-sided Wilcoxon rank sum test with the FDR-adjusted p-values marked in the plots as 0-0.001***, 0.001-0.01**, 0.01-0.05* or NS (non-significant difference).

Then, we analyzed multimorbidity, indicated as the number of concurrent diagnostic categories (Additional file 1: Table S1) among the ASD individuals, revealing that ASD-preterm exhibited a higher likelihood of the higher number of concurrent morbidities compared to ASD-term (Figure 2B, χ² test with *p*<2.2×10^-16^; ORs are recorded in Additional file 1: Table S10). In ASD, preterm sub-groups showed differences in the burden of multimorbidity (Additional file 1: Figure S1B, χ² test with *p*<2.2×10^-16^), in which extremely and moderate preterm subgroups exhibited a significantly higher burden of multimorbidity with >=5 diagnoses compared to late preterm (Additional file 1: Table S11, post-hoc test of χ² test with FDR-adjusted *p*=1.6×10^-5^, 0.02 respectively). For diagnostic categories with more than two specific diagnoses, we found a positive linear correlation between the number of specific diagnoses, i.e., multimorbidity, and the odds ratio of preterm versus term, except for birth-related issues (Additional file 1: Figure S1C). This indicated that preterm ASD individuals tend to have more specific diagnoses within categories as well as across categories than term ASD individuals.

Next, we analyzed quantitative measures for overall behavioral challenges and specific symptom domains. We observed significant differences between ASD-preterm and ASD-term (2-sided Wilcoxon rank sum test with FDR adjustment), although the large sample size may amplify the differences (Figure 2C). ASD-preterm had increased severity of behavioral challenges (CBCL score for 1-5y, *p*=1.4×10^-3^; for 6-18y, *p*=0.0043), developmental coordination disorder (DCDQ final score, *p*=2.5×10^-21^), repetitive behaviors (RBS-R score, *p*=6.7×10^-34^), and social communication skills (SCQ score, *p*=1.4×10^-20^), as well as lower IQ scores (*p*=0.02). Comparing different sub-groups of preterm birth, we found that extremely preterm has the lowest DCDQ final score compared to other stages (2-sided Wilcoxon rank sum test, FDR-adjusted p-values are 0.003, 0.002, and 2.7×10^-5^ when compared to very preterm, moderate preterm, and late preterm, respectively) (Additional file 1: Figure S2).

To complement our analyses within the ASD individuals, we analyzed if there were any differences within preterm birth for the same phenotype measures. The ASD-preterm had more severe outcomes in comparison to non-ASD-preterm with increased severity with lower gestational age (Additional file 1: Figure S3A-B, S4). The developmental diagnostic category had the highest prevalence (72%) in the ASD-preterm group, resulting in 8.8 OR (95% CI 7.9–9.7) when compared to non-ASD-preterm (Additional file 1: Table S12). Also, ASD-preterm group was seven times as likely (OR = 7.0, 95% CI 5.4–9.4) to have five or more multimorbidities than non-ASD-preterm group (Additional file 1: Table S13). For quantitative measures, the ASD-preterm group had statistically significantly higher SCQ final scores compared to the non-ASD-preterm group (2-sided Wilcoxon rank sum test, *p*<2.2×10^-16^) (Additional file 1: Figure S3C).

ASD-preterm and term comparisons within the SSC cohort also showed a higher prevalence of eating and growth problems (OR=2.58 [95% CI 1.79–3.68], and 2.02 [95% CI 1.06–3.58] respectively; Additional file 1: Table S14), and a similar trend towards having more multimorbidity compared to ASD-term (Additional file 1: Figure S5A-B, χ² test *p*=0.045; Additional file 1: Table S15). No statistically significant differences were observed in the quantitative measures (Additional file 1: Figure S5C).

### Genetic variants comparison across ASD and prematurity

To investigate the burden of de novo variants, we analyzed available genome sequencing (GS) and exome sequencing (ES) data from SPARK and SSC. The population analyzed for GS included 310 ASD-preterm, 2,742 ASD-term, and 165 non-ASD-preterm individuals. The ES dataset contained 697 ASD-preterm, 5,747 ASD-term, and 210 non-ASD-preterm individuals. We did not observe any significant difference in DNV event rate or distribution of DNV numbers between ASD-preterm and ASD-term (Figure 3A, Additional file 1: Figure S6A), or between ASD-preterm and non-ASD-preterm derived from GS (Figure 3D, Additional file 1: Figure S6B), even when analyzing only the exonic region (Additional file 1: Figure S7A-B). For the simplex family subset, no differences were observed between preterm and term ASD probands (Additional file 1: Figure S8A). Similarly, no statistically significant differences were found comparing de novo burden from GS in 137 ASD-preterm and 1,313 ASD-term in the SSC (Additional file 1: Figure S7C).

**Figure 3.**
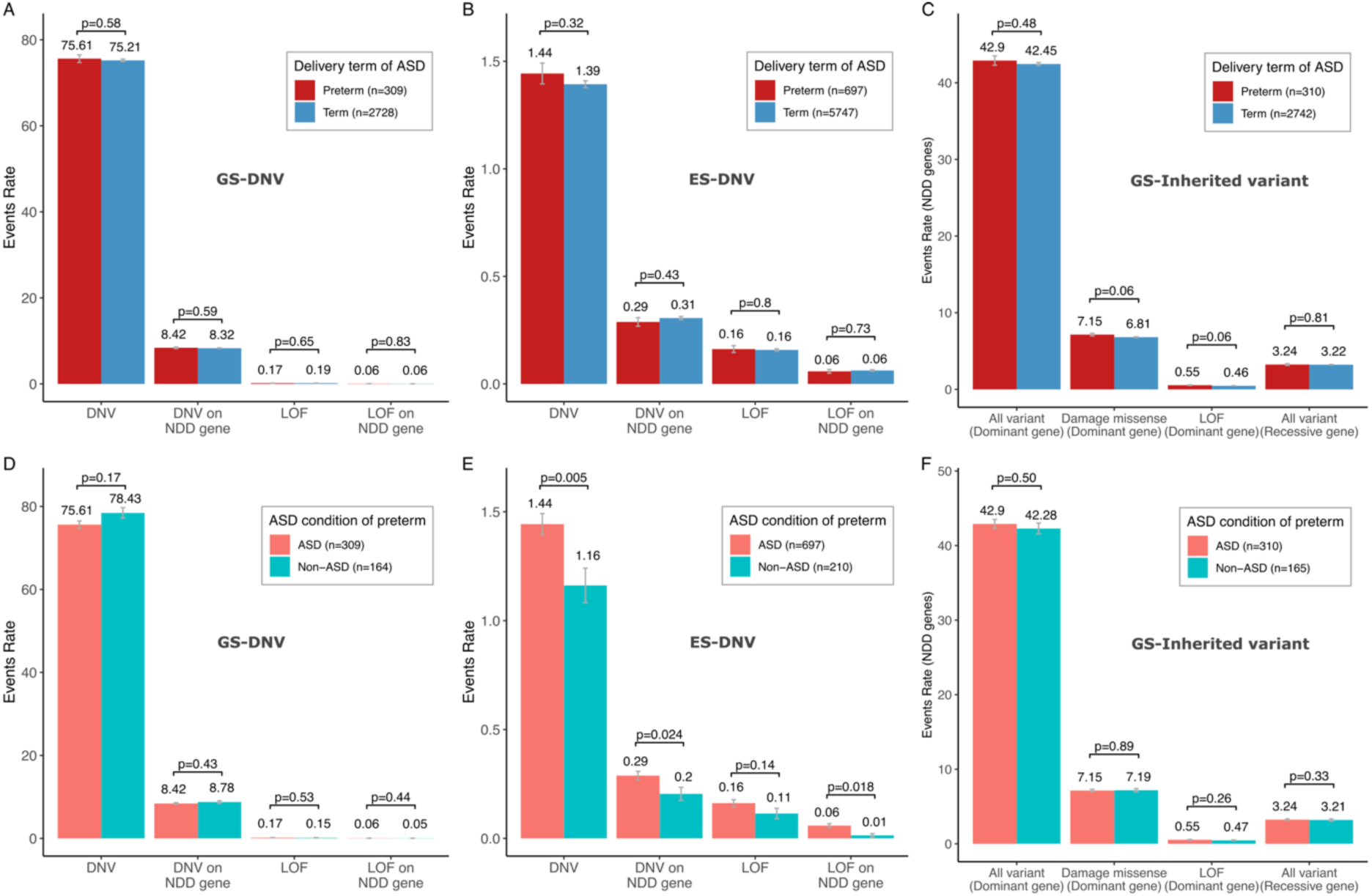
Association between genetic variant burden and subgroups with varying preterm birth and ASD status in the SPARK cohort. In ASD individuals, event rates of rare de novo variants (DNV) identified through genome sequencing (A) and exome sequencing (B), and inherited variants on dominant and recessive NDD genes identified through genome sequencing (C) were calculated. In preterm individuals, event rates of de novo variants identified through genome sequencing (D) and exome sequencing (E), and inherited variants in dominant and recessive NDD genes identified through genome sequencing (F) were calculated. Rare DNVs were filtered with AF<0.001, and rare inherited variants were filtered with AF<0.001 on dominant genes and AF<0.01 on recessive genes. Data are presented as mean values ± standard errors as error bars. The GEE model with Poisson family and sex covariate was used to compute the p-values to assess the differences in variant count between groups.

When analyzing DNV event rates obtained from ES data [8] from SPARK, no statistically significant differences were found between ASD-preterm and ASD-term (Figure 3B, Additional file 1: Figure S6C), even when restricted to probands in simplex families (Additional file 1: Figure S8B). However, ASD-preterm individuals had more exonic DNVs (*p*=0.005), exonic DNVs on NDD genes (*p*=0.024), and LOF affecting NDD genes (*p*=0.018) than non-ASD-preterm (Figure 3E, Additional file 1: Figure S6D). We stratified the ASD-preterm and non-ASD-preterm by gestational age preterm subgroups and observed a similar trend in both the moderate and late preterm groups. However, due to the limited sample size, a statistically significant difference between ASD-preterm and non-ASD-preterm was only detected in the event rate of DNVs in NDD genes in the moderate preterm group (*p*=0.02) and in both overall DNV and DNV in NDD genes in the late preterm group (*p*=0.002 and 0.04, respectively). Interestingly, for extremely to very preterm stages, the event rates of DNV were numerically lower in ASD compared to non-ASD individuals, even though this difference was not statistically significant (Additional file 1: Figure S9).

We also investigated the rates of inherited variants, focusing on those affecting NDD genes and protein-coding regions. From 4,974 individuals with phenotype information, we did not observe statistically significant differences between ASD-preterm and ASD-term nor between ASD-preterm and non-ASD-preterm, although ASD-preterm tend to have a numerically higher rate of rare inherited variants (Figure 3C, 3F, Additional file 1: Figure S6E, S6F). No differences between preterm and term ASD probands in simplex families as well (Additional file 1: Figure S8C).

To further evaluate whether the greater multimorbidity is associated with increased DNV burden, and whether multimorbidity and DNV burden are jointly associated with the increased likelihood of ASD and preterm, we computed GEE models (Additional file 1: Table S16) and found that multimorbidity is positively correlated with GS LOF (*p*=0.037), GS LOF on NDD genes (*p*=5.3×10^-06^) and all types of ES DNV burden (*p*=1.1×10^-^05, 1.9×10^-09^, 6.6×10^-14^ and <2.2×10^-6^ for DNV, LOF, DNV on NDD genes and LOF on NDD genes respectively) across all individuals. Stratified by preterm and ASD status, we observed this positive correlation pattern in ASD-term group for ES DNV (*p*=0.009), ES DNV on NDD genes (*p*=0.013) and LOF on NDD genes (*p*=0.004), as well as in ASD-preterm group for ES LOF on NDD genes (*p*=0.013) (Additional file 1: Figure S10). However, except for GS DNV on NDD genes, there is no interaction between DNV burden and multimorbidity performing on ASD or preterm outcomes.

### ASD Polygenic risk score and association with preterm status

We calculated ASD PRS for individuals in the SPARK cohort using the most comprehensive GWAS on ASD as source data [10]. There was no significant difference in the distribution of PRS between the ASD-preterm and ASD-term groups (even restricted to simplex family probands) nor between ASD-preterm and non-ASD-preterm (Figure 4A). As expected, ASD individuals had higher PRS compared to non-ASD individuals in the whole cohort displaying the usability of the PRS (2-sided Wilcoxon rank sum test, *p*=6.7×10^-13^) (Figure 4A). The PRS explained 9.8% (McFadden Pseudo-r^2^) variance of ASD diagnosis in the SPARK cohort. Additionally, after adjusting for sex and population ancestry (as indicated by principal components [PC]) in a GEE logistic model, we confirmed that there was no independent association between preterm birth and PRS (in ASD population), or between ASD diagnosis and PRS (in preterm population). The statistically non-significant association between preterm birth and PRS (in ASD population) was replicated in the SSC cohort, and same for simplex family probands in SPARK (Additional file 1: Figure S11). Furthermore, we computed a full GEE logistic model for ASD diagnosis within European populations, showing that male sex, preterm birth, and higher PRS were all positively associated with ASD diagnosis (*p*<2×10^-16^, <2×10^-16^ and 2.2×10^-12^, respectively) (Figure 4B). In this model for the SPARK cohort, the predicted probability of an ASD diagnosis was almost 90% for preterm-born males, with the highest PRS (Figure 4C). Then, we included the interaction between preterm status and PRS in the full GEE logistic model, observing a significant association between this interaction and ASD diagnosis (*p*=0.017).

**Figure 4.**
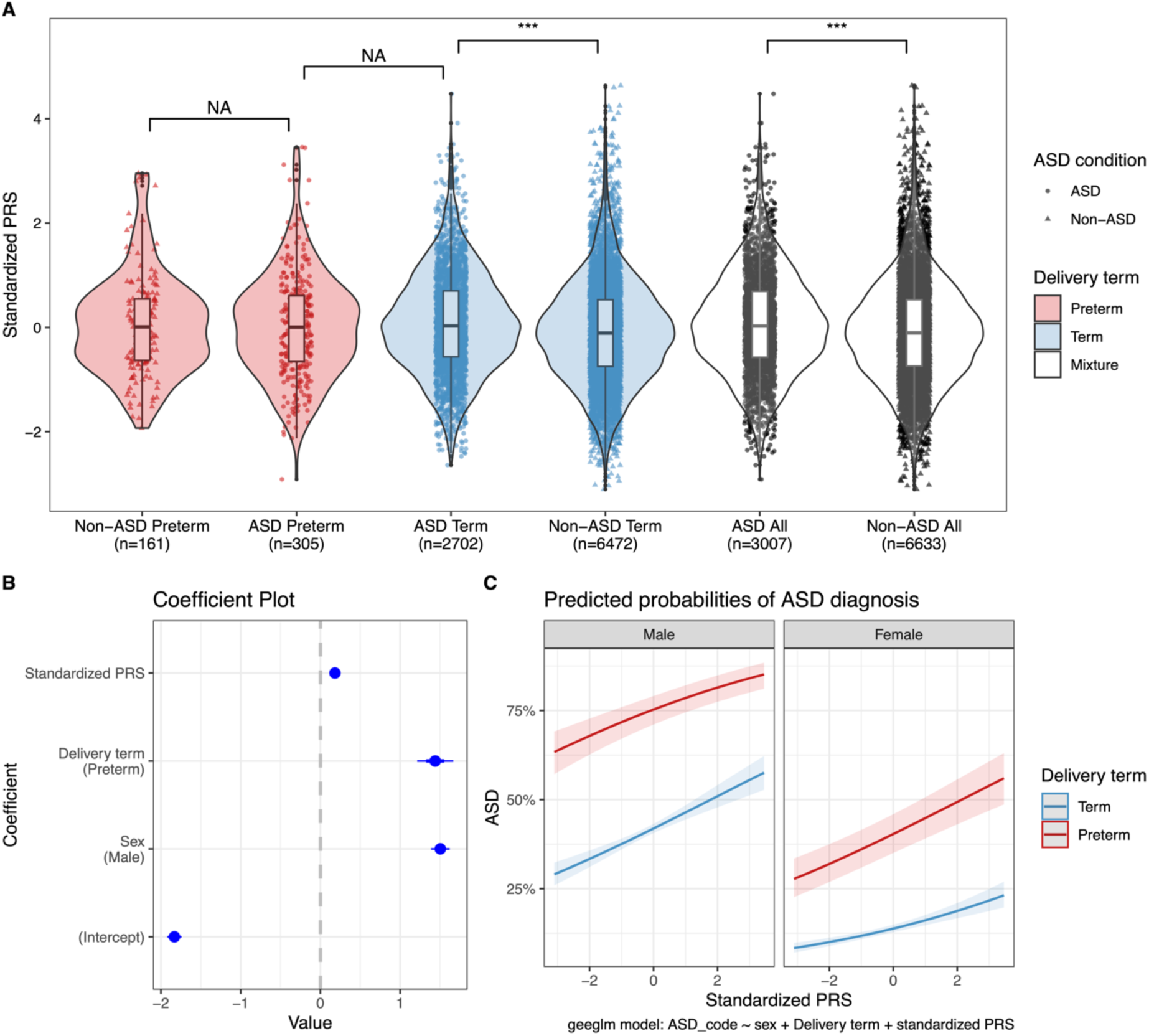
The association of polygenic risk score (PRS) with delivery term and ASD condition in SPARK cohort. A. The distribution of standardized PRS in groups with different delivery terms and ASD diagnosis. Statistical significance was assessed using the 2-sided Wilcoxon rank sum test with the p-value marked in the plots as 0-0.001*** or NS (statistically non-significant difference). B. Coefficient plot for the GEE logistic model [ASD(y/n) ∼ sex(m/f) + Preterm status(y/n) + Standardized PRS] with family ID as a clustering variable, displaying the estimated coefficients for each variable. Positive coefficients suggest an increase in the likelihood of ASD associated with the variable, while negative coefficients indicate a decrease. Error bars represent 95% CI. C. Visualized effect plot of GEE model, which shows average predicted probabilities of ASD diagnosis for specific levels of variables, with color region around the line showing 95% CI.

### Predictive model for ASD within preterm births

Lastly, we investigated the potential of ML models to identify those preterm infants with a high likelihood of ASD from information present at birth by combining clinical and available genetic data.

The model was developed and tested using a study population with preterm individuals classified into ASD (n=279) and non-ASD (n=150). For features used in the prediction model, we also considered Combined Annotation Dependent Depletion (CADD) scores, which assess the potential impact (i.e., deleterious or benign) of genetic variants on the function of genes and were available for most of the DNVs. We applied Recursive Feature Elimination (RFE) and a correlation threshold of 0.7 to select 13 features, including clinical features (sex, condition of birth complications, gestational age, insufficient oxygen at birth) and genomic features (number of several types of variants, CADD scores and standardized ASD-PRS) (Additional file 1: Figure S12A, Table S6). We used three algorithms to train the models (Table 2), of which the XGBoost model exhibited the highest area under the receiver operating characteristic curve (AUROC), at 0.65. The model accurately identified 69% (95% CI 0.644–0.733) ASD diagnoses in the preterm, with a sensitivity of 0.81, specificity of 0.47, and F1-score of 0.77. We also trained a model using the same set of features on all individuals, including both preterm and term (assuming a gestational age of 40 weeks for terms), and despite the sample size (1814 non-ASD and 2880 ASD) being approximately 10 times larger than that of the preterm model, the model performance was comparable (XGBoost with accuracy = 0.69 [95% CI 0.672– 0.699]; AUC = 0.68; F1-score = 0.76; Additional file 1: Table S17).

**Table 2.** Performance metrics of machine learning models used to predict ASD diagnosis in preterm individuals.

| Algorithm | Accuracy | 95% CI | AUC | Sensitivity | Specificity | F1-score |
| --- | --- | --- | --- | --- | --- | --- |
| XGBoost | 0.69 | (0.64, 0.73) | 0.65 | 0.81 | 0.47 | 0.77 |
| Random forest | 0.67 | (0.63, 0.72) | 0.63 | 0.86 | 0.33 | 0.77 |
| SVM | 0.70 | (0.66, 0.75) | 0.62 | 0.84 | 0.46 | 0.79 |

The first three XGBoost models within the 10-fold training (Additional file 1: Figure S12B) were selected to visualize feature effects for the best-performing model for ASD within preterm births. The feature importance varied slightly across the training XGBoost models with sex, PRS, and CADD score being the most important features (Additional file 1: Figure S12C, S12E, S12G). Using SHAP values to characterize the impact of each feature on the model’s output for specific individuals, we found that sex had the highest significant impact on the model’s predictions, whereas being male had a positive impact on the model’s prediction. Furthermore, we demonstrate that lower gestational age, more autosomal exonic DNVs, more dominant inherited variants, more LOF variants, the presence of birth complications, and insufficient oxygen at birth drove the model towards ASD prediction result (Additional file 1: Figure S12D, S12F, S12H).

## Discussion

Here, we conducted a comprehensive analysis of phenotypic differences using larger cohorts, as well as genotypic differences, which have been explored in only a few studies among preterm and term-birth ASD individuals. We conclude that preterm-born ASD individuals have more diagnoses across different categories and a number of co-occurring diagnoses but similar genetic landscapes when investigating sequence-level rare DNVs and inherited variants as well as a polygenic load for ASD compared with ASD-term. Our analysis of preterm individuals with and without ASD showed similar results for the phenotype comparisons but inconsistent findings for the genetic burden. The largest de novo dataset derived from ES showed that the ASD-preterm had a higher exonic DNV event rate than the non-ASD-preterm; however, we did not validate this finding in the de novo dataset from GS. Additionally, the male with preterm status and higher polygenic load faces a higher likelihood of ASD when considering these features together. Furthermore, our ML model demonstrated potential for predicting ASD diagnosis in preterm children by integrating phenotype and genetic information. Our results provide evidence that genetic factors play a role in emerging ASD in preterm birth, but the environmental stressor of being preterm potentially contributes to the severity and multimorbidity.

Previous research has reported numerous but inconsistent findings regarding phenotypic disparities between ASD preterm and term individuals [20,22], while limited research has focused on investigating the genetic link of ASD in prematurity. Unlike most phenotypic comparisons that concentrate on specific diagnostic outcomes [21,22], we first grouped the various conditions into nine broader diagnostic categories. Our results indicate that children with both preterm birth and ASD exhibit a higher prevalence of diagnoses within these categories and a higher rate of multimorbidity across different diagnostic categories. To note, we did not replicate all the observed associations in SSC cohort, which had mostly late preterm category for the GA and more restrictive recruitment criteria, which could explain the lack of replication. Previous studies have found that both preterm birth and ASD are associated with adverse symptoms. For instance, preterm infants are independently inherently prone to multimorbidity and severe health complications affecting multiple organs and systems [17,53,54], such as visual and auditory impairments [55], epilepsy [56], ADHD [18], and other psychiatric disorders [19]. Additionally, individuals with ASD experience a higher burden of co-occurring medical conditions [57]. Our study further supports the hypothesis that preterm acts as an environmental liability factor, potentially influencing some of the heterogeneity and higher comorbidity rates observed in ASD.

After stratifying preterm based on gestational age, we observed that those born with lower gestational age tend to have more severe outcomes, which is in line with the dose-effect reported in prematurity, where the likelihood of developmental issues increases with decreasing gestational age [54]. This effect is also reflected in the potentially increasing complexity of multimorbidity among groups with lower gestational age [17]. Additionally, we showed significantly more severe symptom levels, as measured by different standardized questionnaires and cognitive tests, in ASD-preterm, consistent with previous studies as well as general research comparing preterm and term birth [18,58–60]. It is important to note that with large sample sizes, even very small differences can become statistically significant. In SSC cohort, with over 90% of preterm individuals being late preterm, thus replication of results was limited. Therefore, the results should be interpreted with caution especially for clinical utility.

In idiopathic ASD, heritability is estimated to be approximately 80% [6], but in preterm born, environmental factors account for 60% of the variation in gestational age [61]. Our findings suggest that genetic factors underlie, at least partly, the ASD diagnosis even in preterm, but that the complex phenotypic presentation, including multimorbidity, could be due to the environmental stressor of being preterm. Specifically, we did not observe significant differences in DNV numbers between preterm and term ASD individuals. We did observe suggestive evidence that DNV burden could be higher in ASD-preterm compared with non-ASD-preterm, however, the finding was inconsistent in the validation cohort with a smaller sample size and after stratifying by preterm stages. We speculate that this may be indicative of distinct underlying genetic mechanisms for ASD across different preterm sub-groups. Across preterm, in light of previous reports showing that ASD has a higher DNV burden than non-ASD [7], we suspect that some DNVs specifically affect ASD liability but not prematurity, or some DNVs could be liability factors for both ASD and prematurity. Limited research indicated a higher DNV burden in overall preterm newborn genomes and primarily in genes related to embryonic brain development; however, the study did not consider ASD or another behavioral diagnosis in preterm infants [23], especially ASD also susceptibility to brain development-related genes [62]. For the lack of differences in DNV burden observed between preterm and term in ASD, one hypothesis is that some DNVs contributing to ASD have global effects on overall physiology and also contribute to prematurity. In this situation, those DNVs could possibly increase the likelihood of both preterm birth and ASD with multimorbidity. Furthermore, it remains to be clarified to what extent prematurity acts as an environmental liability factor to ASD when counting in genetic background.

Although GWAS studies of prematurity have identified variations in maternal and fetal genes separately [63,64], few have examined the impact of rare inherited variants. Our study did not find a difference in the burden of rare inherited variants between ASD-preterm and ASD-term individuals. This can be partially explained by the fact that the maternal genome influences prematurity more than the fetal genome [15]. Additionally, we have perhaps overlooked shared genetic factors that potentially contribute to both preterm birth and neurodevelopment when we analyze individuals with either ASD or prematurity. For example, variants at *AGTR2* and *ADCY5* genes in mothers have been associated with gestational duration and preterm birth, and infants show around half of effect size by sharing one of the maternal allele by descent [65]. Notably, these two genes are also known ASD-associated genes [40].

Although we did not find an overall association between ASD PRS and prematurity, we show intriguing findings that in the SPARK cohort, those with the highest PRS could have a higher likelihood of ASD, especially in preterm infants and boys. Even after including the interaction between preterm status and PRS, these features maintained a significant association with ASD likelihood, and the interaction itself was significantly associated with ASD diagnosis. The lowest PRS in preterm males still reached a 75% predicted probability of ASD, emphasizing the prematurity effect on ASD liability and the greater susceptibility in males, while also indicating a potential sample bias towards ASD in our analysis. Again, these findings need validation in the general population, especially as a prior study by Cullen et al. found no evidence of an interaction effect between ASD polygenic score and gestational age at birth on cognition [16]. However, it is important to note that the cognitive difficulty they measured is only one of the outcomes that do not imply an ASD diagnosis, and the model they used also included socio-economic status as a covariate.

In addition to genetic factors, widespread alterations in brain development associated with preterm infants may contribute to the increase in ASD likelihood. Previous studies have indicated that reduced structural brain asymmetry and poor brain development during neonatal life may increase the liability of ASD in preterm infants [66,67]. Even in preterm children exhibiting similar ASD traits during childhood, distinct etiological trajectories have been observed involving variations in neonatal cerebellar volume and developmental delay [68].

Not all preterm infants develop ASD [69], still we demonstrate that when genetic factors are combined with the environmental risk of preterm birth, preterm children face a highly elevated likelihood of ASD diagnosis. Recognizing the limitations of traditional statistical models in capturing nonlinear interactions between features, we developed an ML model to predict ASD diagnosis in preterm children at birth. Unlike previous ASD prediction models that rely on developmental trajectories or typical characteristics collected as children grow [26,30,70], our ML model utilized only information available at birth, integrating phenotype and genetic information. Moreover, most previous models are based on the general population [26,70], limiting their applicability to preterm infants. However, it is necessary to build prediction models tailored specifically for preterm infants due to the heterogeneity of ASD phenotypes [1], and preterm ASD children may exhibit specific phenotypes compared to term ASD children [19]. Although our ML model did not achieve significantly higher performance, achieving 69% accuracy with a small sample size and few features demonstrates the feasibility and efficacy of integrating phenotype and genetic information for ASD prediction. There is still substantial room for improvement in the model performance. Increasing the sample size and adding more features associated with preterm birth and ASD (e.g., maternal age, prenatal exposure, and fetal birth weight) would benefit the prediction model construction. Other features like intubation in the delivery room, family language, parental education, Infection, and ventilation have also been found to have predictive ability for cognitive outcomes in very preterm infants [30]. It is important to note that we cannot identify the causal relationships between features selected by the model and ASD diagnosis. Still, we suggest that these features could potentially enhance prediction models in the future.

Our study has several limitations. Firstly, two cohorts are ASD-focused, and the non-ASD groups are mainly relatives of ASD probands, thus introducing sample bias to ASD and potentially underestimating genetic differences between the groups. Given the high heritability estimation in ASD, siblings with a closer relationship with ASD have a higher relative risk ratio for ASD [4]. Secondly, we did not stratify the analyses by sex due to the limited sample size, potentially overlooking sex-specific differences. Thirdly, we focused here only on the sequence level variation, incorporating more types of genetic variations will be the next step. Additionally, our exploration of genetic factors primarily focused on average population-level associations and NDD genes, potentially overlooking genetic effects beyond the currently known ASD-associated genes and variants that may contribute to the elevated likelihood of ASD in preterm children. Previous studies have pointed out the genetic association between preterm and ASD, such as common genetic variants linking abnormalities in the gut-brain axis with both conditions [71]. Finally, the lack of comparability of SSC with the SPARK cohort may reflect both reduced statistical power and cohort differences. The SPARK cohort collected participants with different family structures and highly heterogeneous clinical severity. While the SSC cohort with a smaller sample size excluded probands with conditions might reduce the validity of diagnoses (e.g., known genetic syndromes, significant perinatal complications, and low mental age). In particular, the distribution of gestational age differs in two cohorts, and more than 90% of preterm in SSC are late preterm. Also, the sample size is low for some of the analyzed groups despite similar percentages of preterm in ASD in the two cohorts. Further validation in larger preterm cohorts or general population-based samples is a key future direction. We believe that combining genetic features and more detailed phenotypic information will help to explain further why some preterm children have ASD while others do not.

## Conclusion

In conclusion, we demonstrate that ASD genetic liability is similar in ASD-term and ASD-preterm, suggesting that even within preterm, genetic factors play an important role in etiology. Our study did not find evidence of a link between genetic factors and preterm birth in ASD. However, our findings suggest that preterm birth would exacerbate the severity of outcomes in ASD individuals, and this difference may be driven more by environmental factors. As we observed some differences in the rate of ES DNV in preterm individuals compared between ASD and non-ASD, we only suggest that genetic factors may increase the likelihood of a preterm child getting an ASD diagnosis and the diagnosis is not modified by the interaction between multimorbidity and DNV burden. Through the development of our ML model, we demonstrate that integrating phenotype and genetic information is feasible and holds promise for the early prediction of ASD in preterm children at birth. Our study provides insights into the phenotypic characteristics of ASD preterm individuals. We suggest that health screening for preterm birth infants should incorporate the collection of genetic data, as it better supports early clinical identification of ASD and can aid in the guidance of early intervention strategies.

## Data Availability

The data that support the findings of this study are available from the Simons Foundation Autism Research Initiative (SFARI, https://www.sfari.org/resource/sfaribase) but restrictions apply to the availability of these data, which were used under license for the current study, and so are not publicly available. Data are however available from the authors upon reasonable request and with permission of SFARI.

## List of abbreviations

ASD: Autism Spectrum Disorder
SPARK: Simons Foundation Powering Autism Research for Knowledge
SSC: Simons Simplex Collection
GS: Genome sequencing
ES: Exome sequencing
GEE: Generalized Estimating Equations
PRS: Polygenic Risk Score
DNV: De Novo Variants
CNV: Copy number variation
GWAS: Genome-Wide Association Studies
NDD genes: Neurodevelopmental Disorders-Related Genes
ML: Machine Learning
CBCL: Child Behavior Checklist
DCDQ: Developmental Coordination Disorder Questionnaire
RBS-R: Repetitive Behavior Scale-Revised
SCQ: Social Communication Questionnaire
Fsiq: Full-Scale Intelligence Quotient
AB: Allele Balance
LOF: Loss-of-Function
CADD: Combined Annotation Dependent Depletion
PCA: Principal Component Analysis
PC: Principal Components
CI: Confidence Interval
RFE: Recursive Feature Elimination
XGBoost: Extreme Gradient Boosting
RF: Random Forest
SVM: Linear Support Vector Machine
AUROC: Area Under the Receiver Operating Characteristic Curve
SHAP: SHapley Additive exPlanations
GA: Gestational Age at Birth
OR: Odds Ratio
FDR: False Discovery Rate

## Additional files

Additional file 1: Supplementary Figures S1–S12 and Supplementary Tables S1–S17. (PDF 4,198 kb)

Additional file 2: Neurodevelopmental disorder-related gene list used in this study. (XLS 65 kb)

## Declarations

### Ethics approval and consent to participate

Ethical approval for the data collection and informed consent were obtained from the participants within the SPARK and SSC projects. The Swedish Ethical Committee approved this study and data analysis in Sweden (dnr 2020-00400).

### Consent for publication

Not applicable.

### Availability of data and materials

The data that support the findings of this study are available from the Simons Foundation Autism Research Initiative (SFARI, https://www.sfari.org/resource/sfaribase) but restrictions apply to the availability of these data, which were used under license for the current study, and so are not publicly available. Data are however available from the authors upon reasonable request and with permission of SFARI. The R scripts used to perform the main analysis reported in this manuscript are available on GitHub (https://github.com/Tammimies-Lab/AutismPreterm_Zhang).

### Authors’ contributions

Y.Z. and K.T. designed the study and planned the analyses. Y.Z. performed the analyses. Y.A. provided support for the analysis. S.S., U.Å, and K.T. provided supervision and support for the analysis. Y.Z. wrote the first draft with feedback from K.T. All authors provided critical feedback and helped shape the research, analysis, and the final manuscript.

## Acknowledgements

We thank all the participants of SPARK and SSC cohorts and Simons Foundation for access of the data. The computations were performed using resources provided by the National Academic Infrastructure for Supercomputing in Sweden (NAISS) through Uppsala Multidisciplinary Center for Advanced Computational Science (UPPMAX). Furthermore, research reported in this publication was supported by the Eunice Kennedy Shriver National Institute of Child Health & Human Development of the National Institutes of Health under Award Number R01HD113669. The content is solely the responsibility of the authors and does not necessarily represent the official views of the National Institutes of Health.

