## Supplementary materials for "Prematurity and Genetic Liability for Autism Spectrum Disorder"

### Additional file 1

Supplementary Figures S1-12
Supplementary Tables S1-17

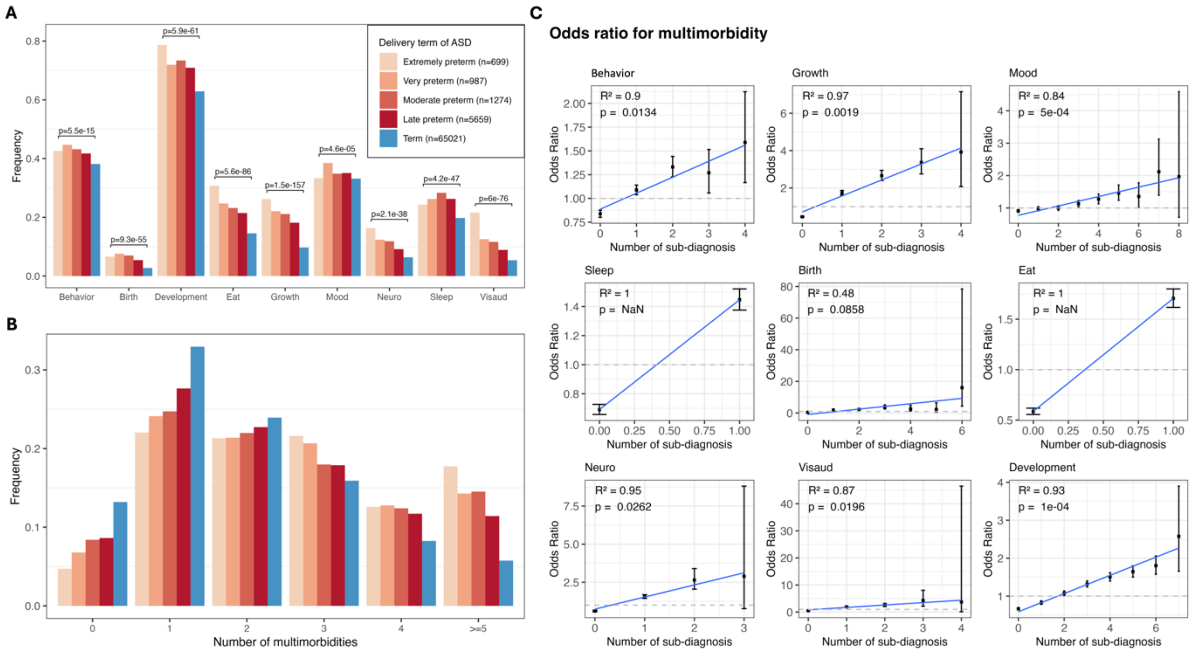

**Figure S1. The phenotype comparison between different preterm stages within ASD regardless of genotype (for the SPARK cohort).** Color bars are the same across three panels and shown in panel A. A. Prevalence of diagnostic categories. Significance was assessed using $\chi^2$ test with FDR-adjusted p-value (details of diagnostic categories are described in Table S1). B. Distribution of the number of multimorbidities (number of diagnostic categories detailed in table S1). The bar color and sample size for each group are the same as in A. The significance of differences in prevalence between different preterm stages is recorded in Table S9. C. The odds ratio (OR) with 95% CI for the frequency of the number of multimorbidities in each diagnostic category. ORs are given among ASD individuals born preterm vs term. The Pearson test was used to assess the $R^2$ and p-value of linear regression.

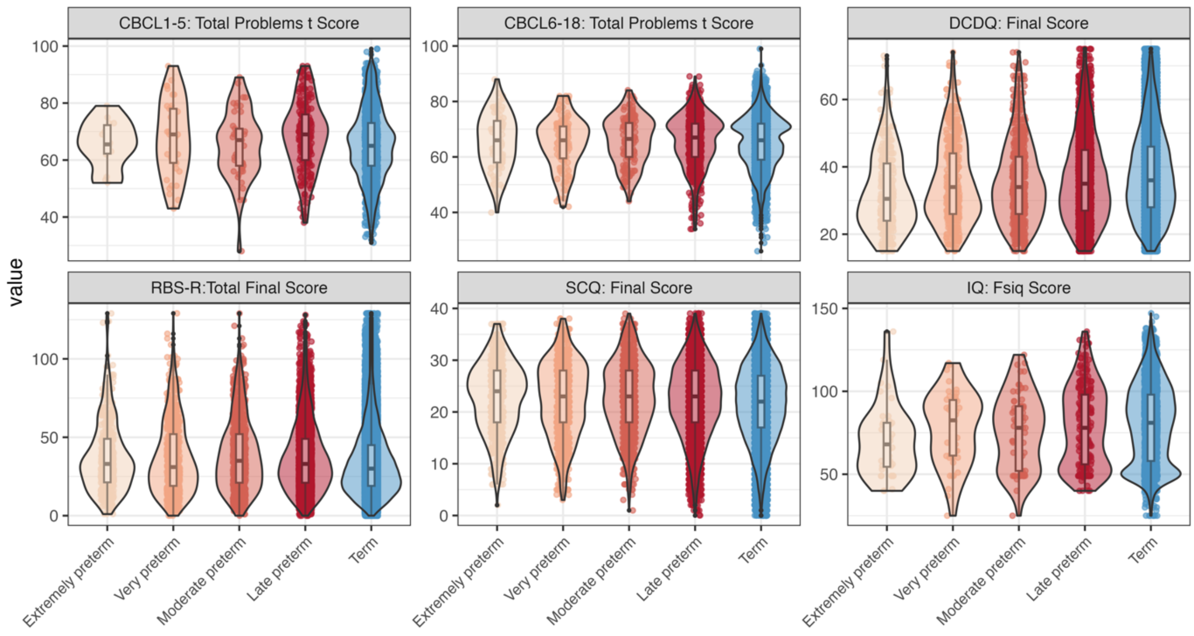

**Figure S2. Differences in quantitative measures among ASD individuals in different preterm stages and term (for SPARK cohort).** Measures include Child Behavior Checklist (CBCL) t score for 1 to 5 and 6 to 18 years of age, Developmental Coordination Disorder Questionnaire (DCDQ), Repetitive Behavior Scale-Revised (RBS-R) score, Social Communication Questionnaire (SCQ), and Full-Scale IQ (Fsiq). Score distributions of different stages were compared using Kruskal-Wallis rank sum test, except CBCL6-18 ($p$=0.089), IQ ($p$=0.066), others have $p$<0.05. In the pairwise Wilcoxon rank sum test, extremely preterm have lower DCDQ final score than very preterm ($p$=0.003), moderate preterm ($p$=0.002), and late preterm ($p=2.7\times10^{-5}$) born, with other pairs having $p$>0.05.

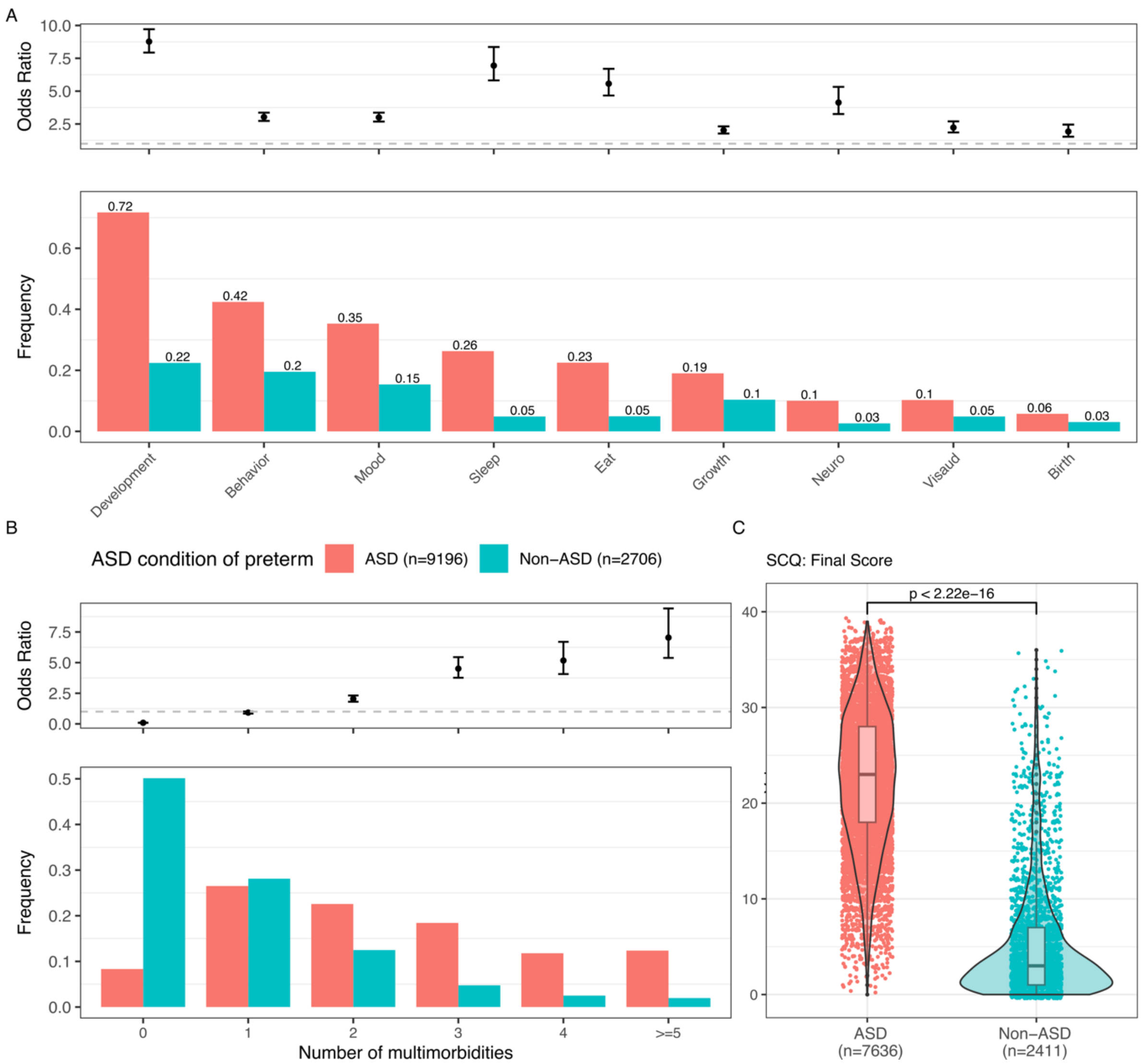

**Figure S3. The phenotype comparison between ASD and non-ASD within preterm individuals in SPARK cohort.** Color bars are the same across three panels and shown at the top of panel B. A. Prevalence and odds ratio with 95% confidence interval (CI) of diagnostic categories. The detailed outcomes in each diagnostic category are described in Table S1. The exact prevalence values are labeled on the top of the bars. ORs are given among ASD individuals vs non-ASD individuals, recorded in Table S12. B. Distribution of the number of multimorbidities, defined as the total number of diagnostic categories that appeared in individuals. ORs with 95% CI are given among ASD individuals vs non-ASD individuals when focusing on each multimorbidity number and recorded in Table S13. C. Differences in Social Communication Questionnaire (SCQ) among preterm individuals with ASD and without ASD. Significance was assessed using the 2-sided Wilcoxon signed-rank test with the p-value marked in the plot as 0-0.001***.

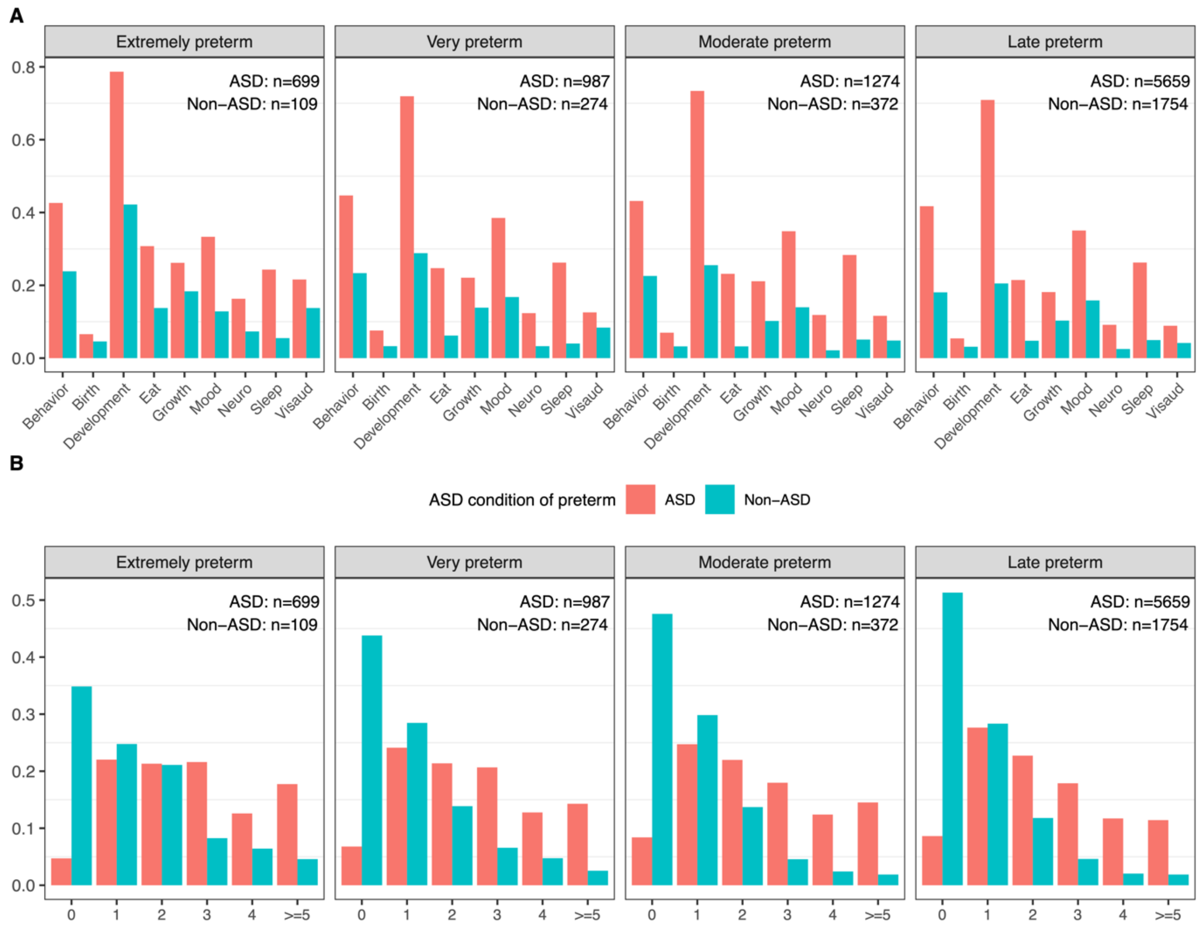

**Figure S4. The phenotype comparison between ASD and non-ASD within different preterm stages in SPARK cohort.** A. Prevalence of diagnosis. B. Distribution of the number of multimorbidities. The sample size of each subgroup is labeled in each panel.

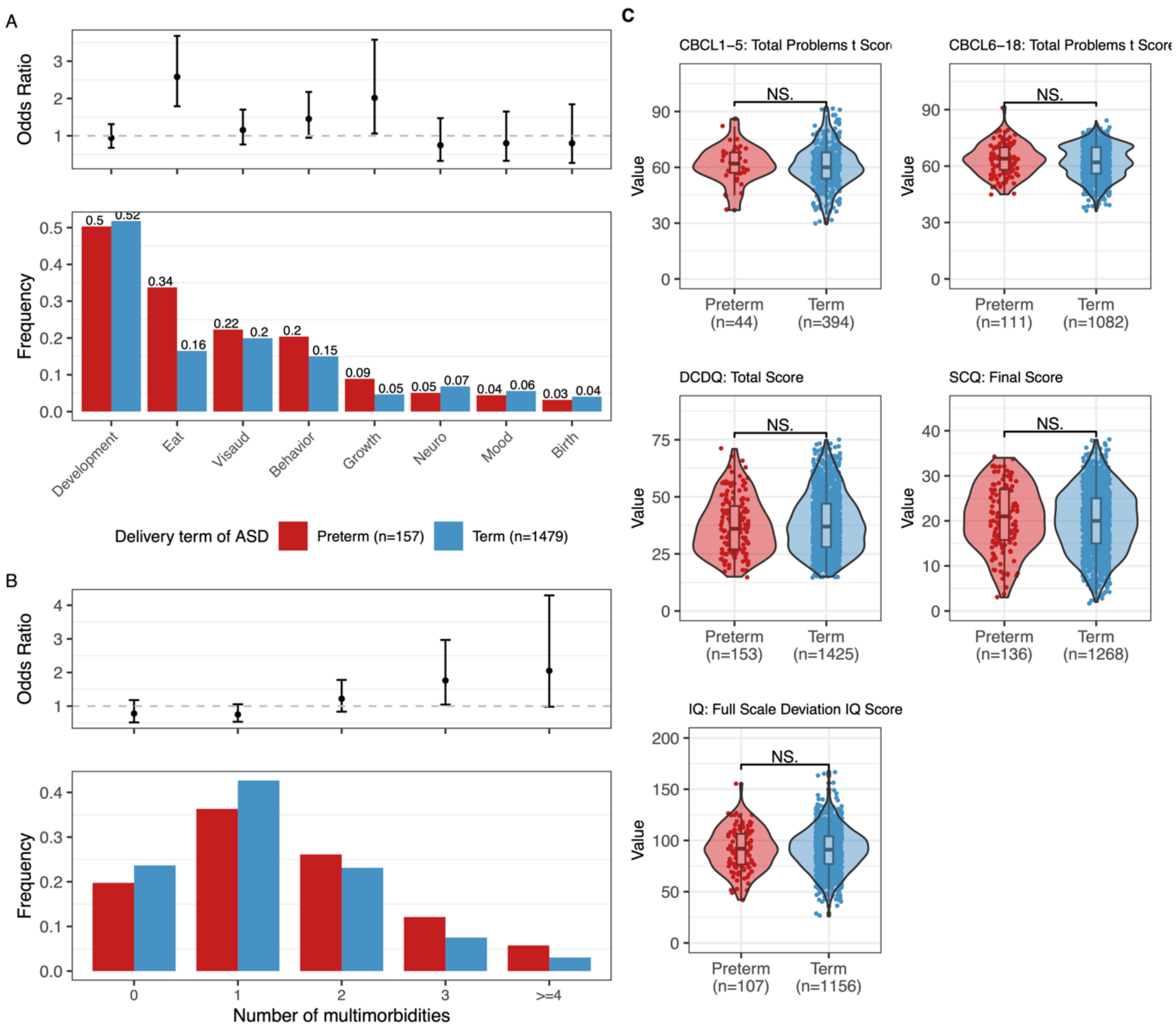

**Figure S5. The phenotype comparison between preterm and term within ASD in the SSC cohort.** A. Prevalence and odds ratio with 95% confidence interval (CI) of diagnostic categories. The detailed outcomes in each diagnostic category are described in Table S8. The exact prevalence values are labeled on the top of the bars. ORs are given among ASD individuals born preterm vs term and recorded in Table S14. B. Distribution of the number of multimorbidities, defined as the total number of diagnostic categories that appeared in individuals. ORs with 95% CI are given among ASD individuals born preterm vs term when focusing on each multimorbidity number and recorded in Table S15. C. Differences in Child Behavior Checklist (CBCL) t score for 1 to 5 and 6 to 18 years of age, Developmental Coordination Disorder Questionnaire (DCDQ), Social Communication Questionnaire (SCQ), and Full-Scale IQ among ASD individuals born preterm and term. Significance was assessed using the 2-sided Wilcoxon signed-rank test with the p-value marked in the plots as NS (non-significant difference).

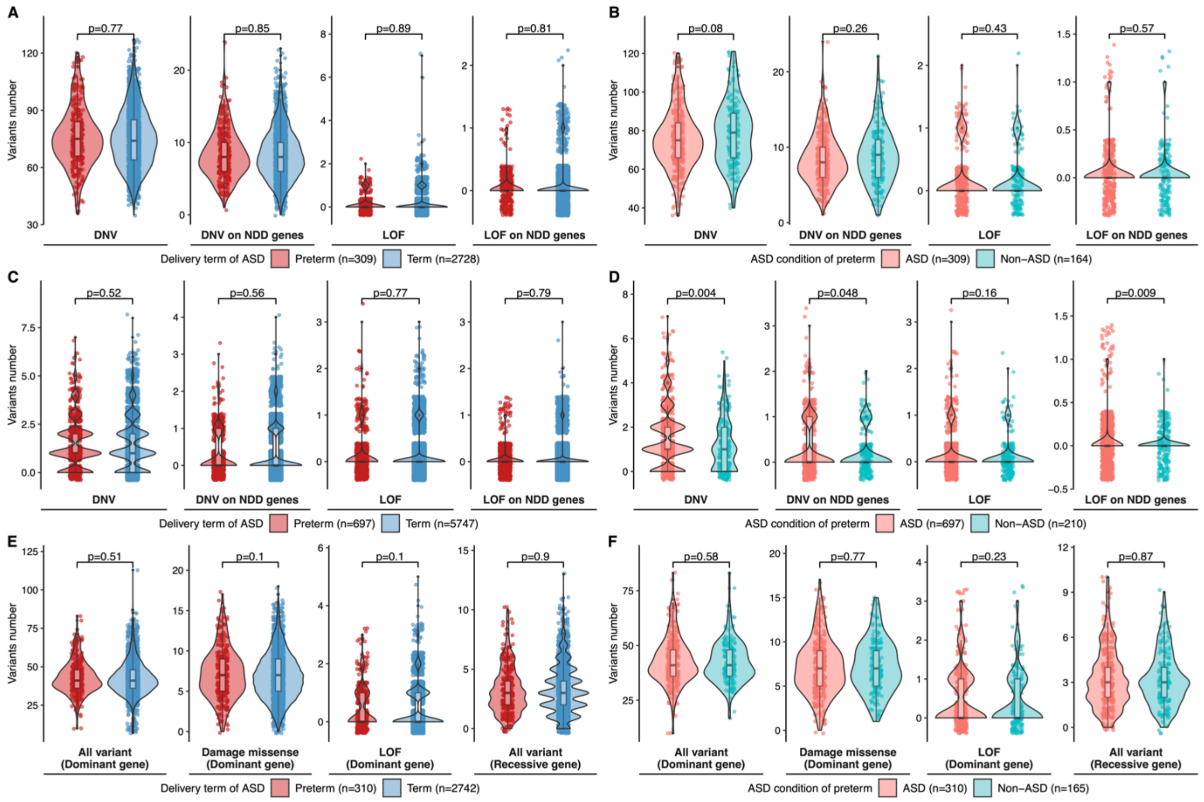

**Figure S6. The distribution of genetic variants number in SPARK cohort.** In ASD individuals, the number of de novo variants identified through whole genome sequencing (A) and whole exome sequencing (C), and inherited variants on dominant and recessive NDD genes identified through whole genome sequencing (E) for each individual were shown. In preterm individuals, the number of de novo variants identified through whole genome sequencing (B) and whole exome sequencing (D), and inherited variants on dominant and recessive NDD genes identified through whole genome sequencing (F) for each individual were shown. Differences between groups were assessed using the 2-sided Wilcoxon signed-rank test.

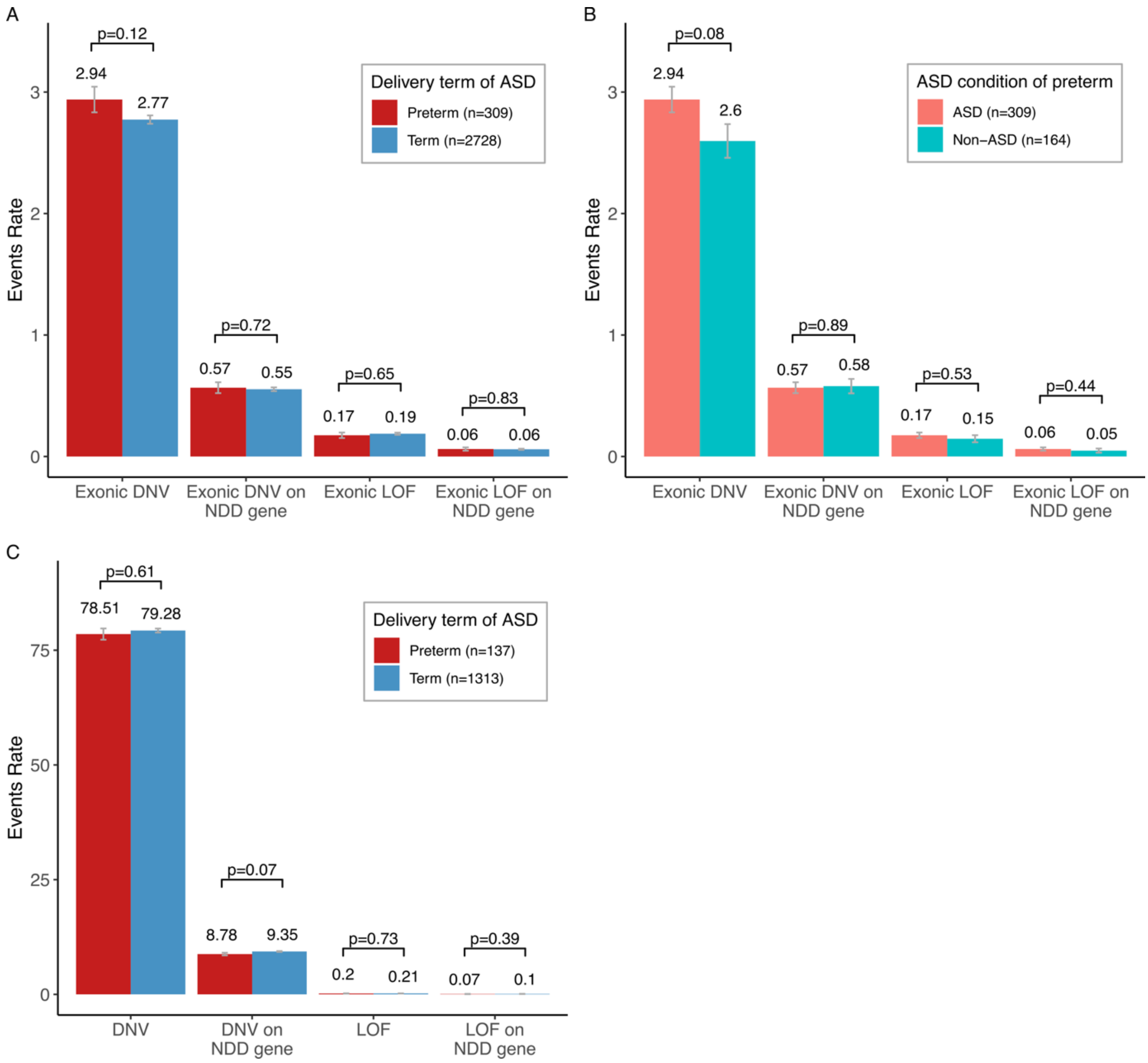

**Figure S7. Association between genetic variant burden and subgroups with varying preterm birth and ASD status. In SPARK,** for de novo variants found on exonic regions in GS, event rates were compared between ASD-preterm and ASD-term (A), and between ASD-preterm and Non-ASD-preterm (B). C. The event rates of de novo variants were compared between preterm and term within ASD individuals in the SSC cohort. Data are presented as mean values ± standard errors as error bars. GEE model with Poisson family and sex covariate was used to compute the p-value to assess the differences in variant count between groups.

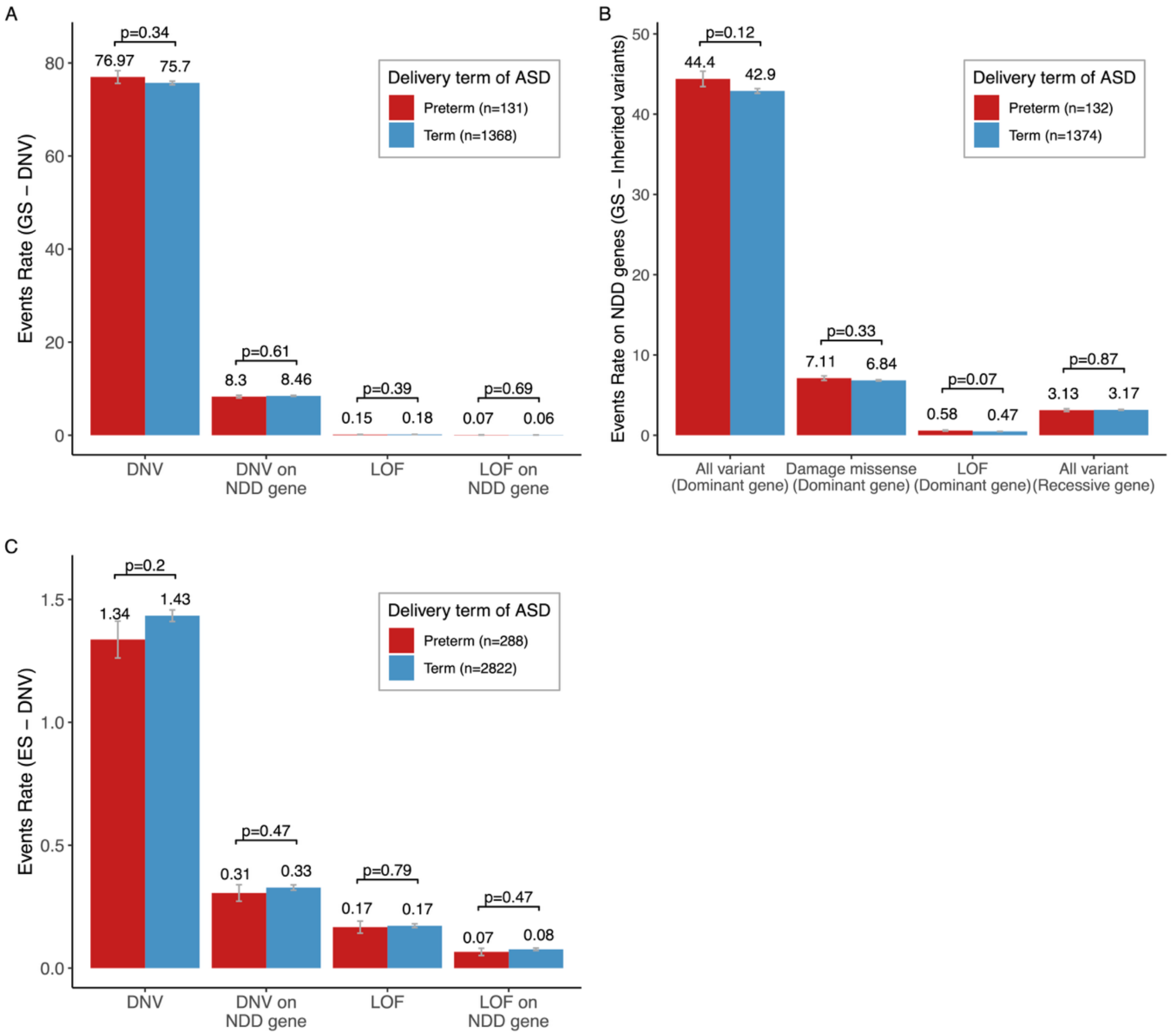

**Figure S8. Association between genetic variant burden and subgroups with varying preterm birth status for probands from simplex families in SPARK.** Event rates of rare de novo variants (DNV) and inherited variants on dominant and recessive NDD genes (B) identified through genome sequencing, and DNV identified through exome sequencing (C) were calculated. Data are presented as mean values ± standard errors as error bars. GEE model with Poisson family and sex covariate was used to compute the p-value to assess the differences in variant count between groups.

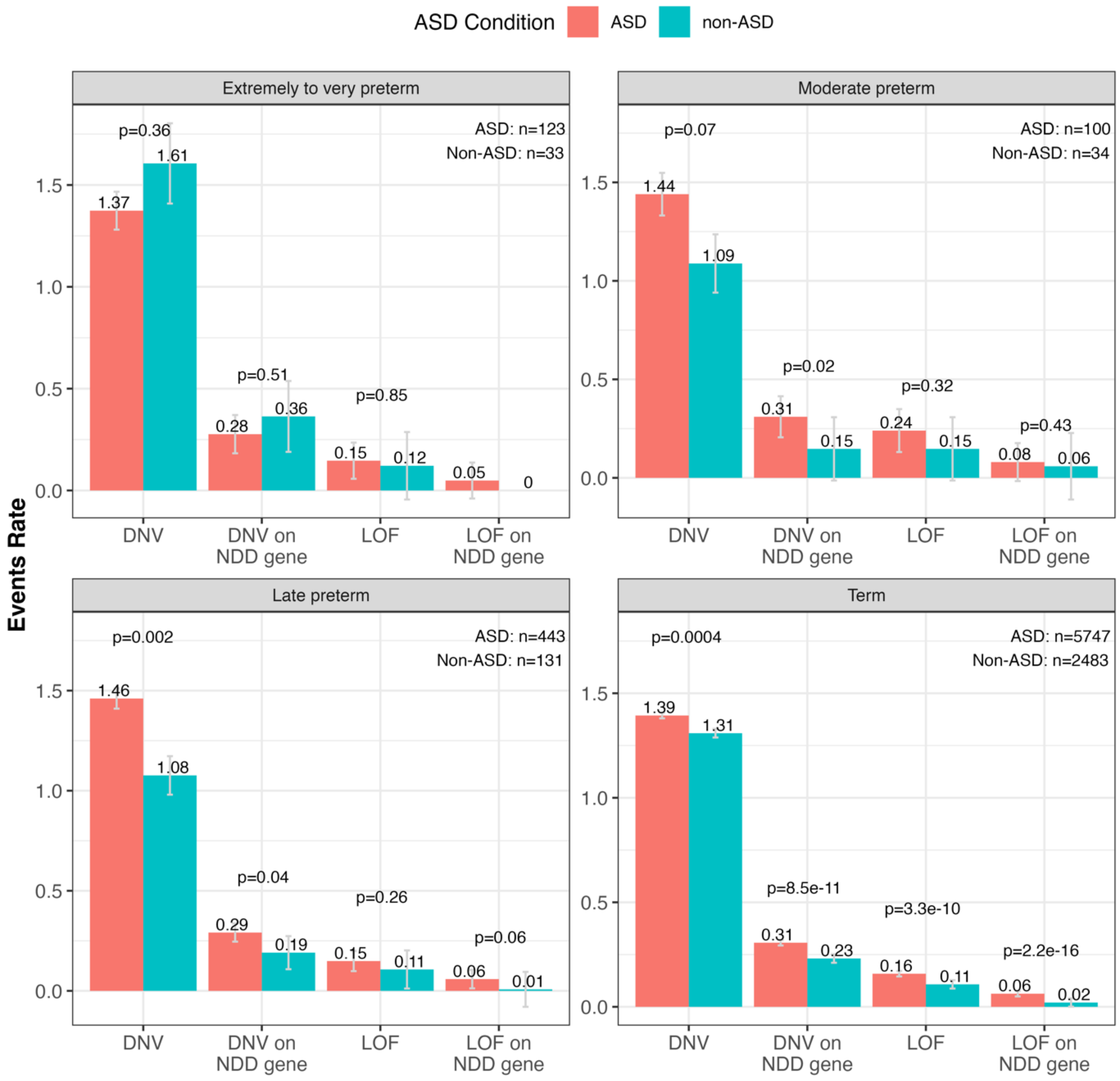

**Figure S9.** The event rates of de novo variants identified in WES were compared between ASD and non-ASD in SPARK individuals in different preterm stages and term. Data are presented as mean values ± standard errors as error bars. GEE model with Poisson family and sex covariate was used to compute the p-value to assess the differences in variant count between groups.

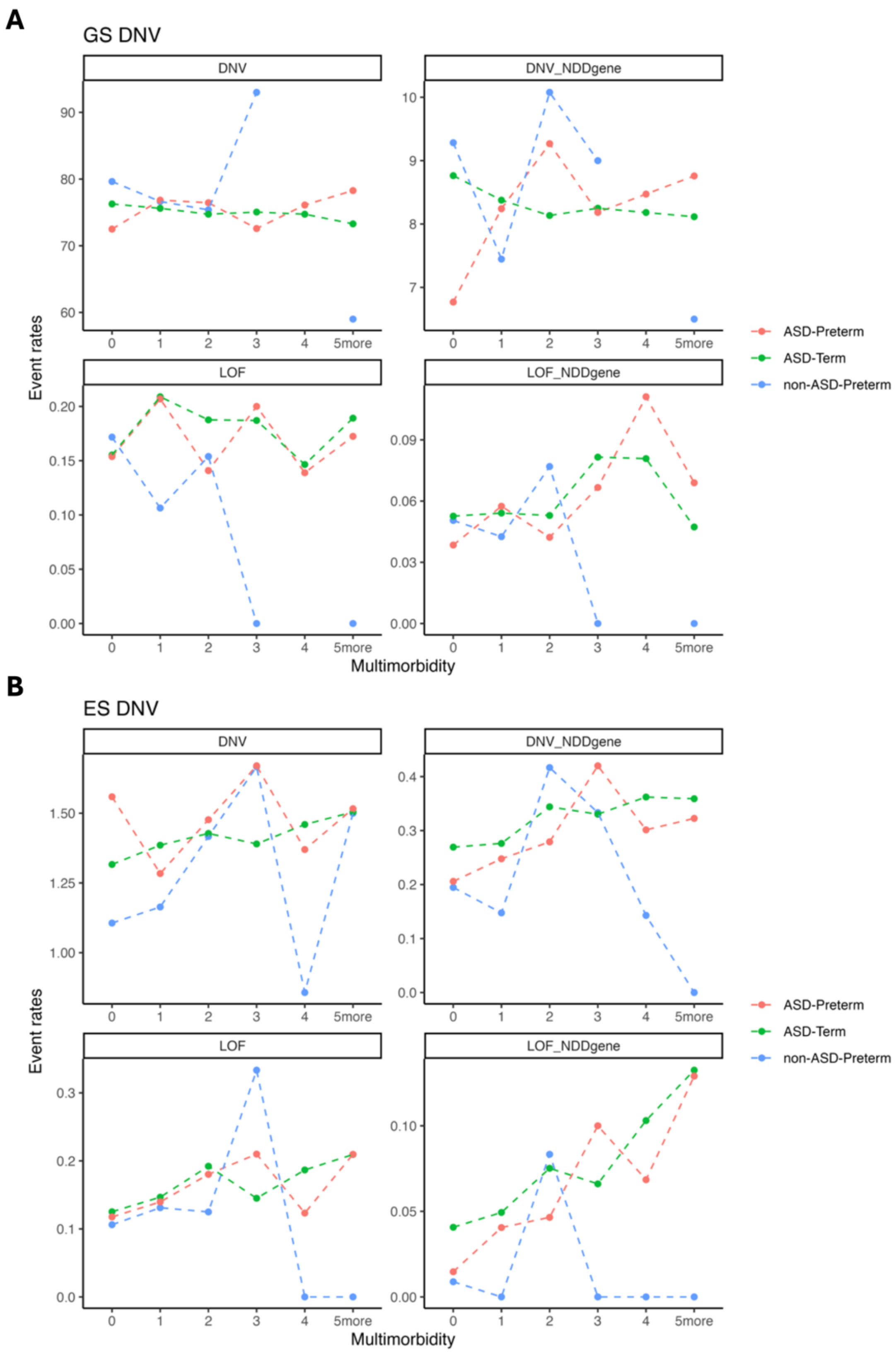

**Figure S10. DNV event rates across multimorbidity categories in different ASD and preterm status (SPARK cohort).** Variants identified in GS (A) and ES (B) are grouped in DNV, DNV in NDD genes (DNV_NDDgene), loss-of-function (LOF) variants, and LOF variants in NDD genes (LOF_NDDgene).

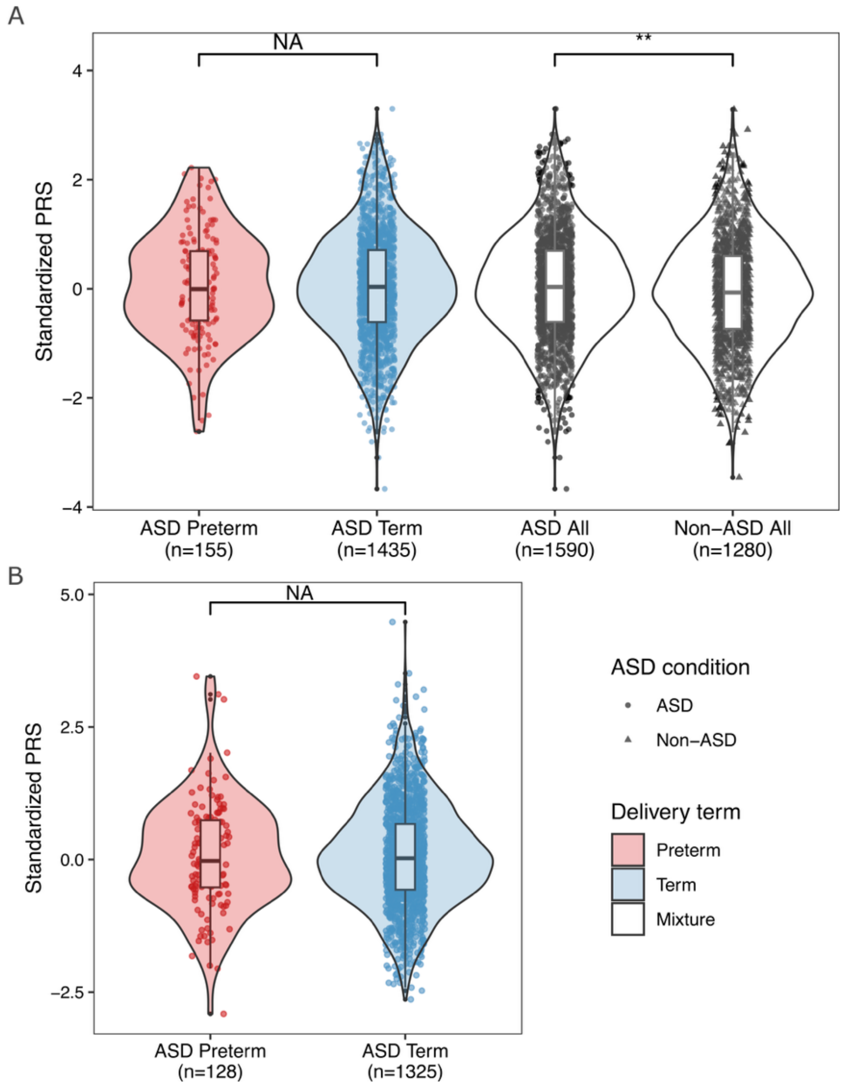

**Figure S11. The distribution of standardized PRS in SSC cohort individuals (A) and SPARK simplex family probands (B) with different prematurity and ASD status.** Significance was assessed using the 2-sided Wilcoxon signed-rank test with the p-value marked in the plots as 0-0.001***, 0.001-0.01**, 0.01-0.05* or NS (non-significant difference).

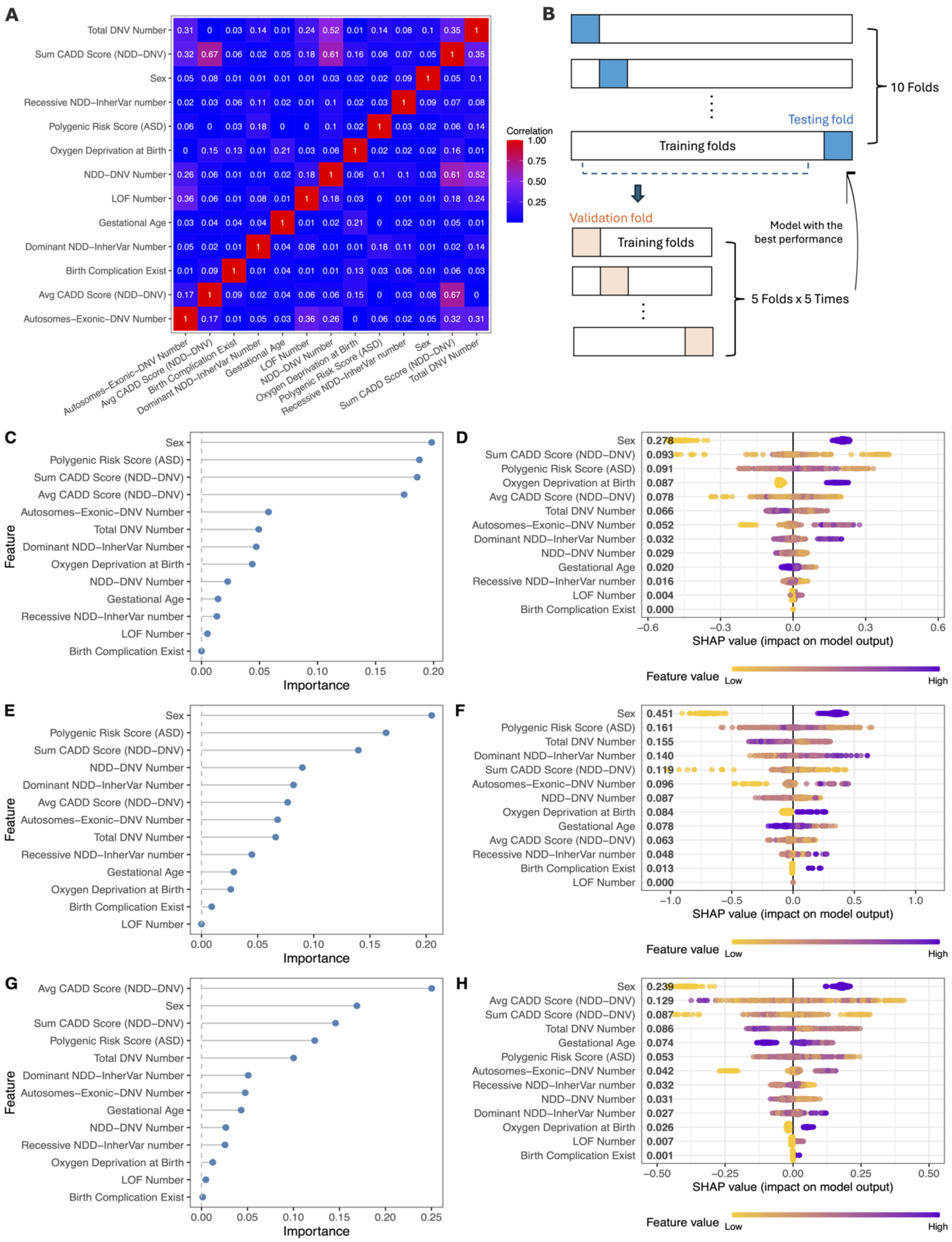

**Figure S12. The design and evaluation of machine learning (ML) model.** A. The correlation between features used in the ML model. B. The model training design. Feature importance (C, E, G) and SHAP values (D, F, H) for ML models trained with the XGBoost algorithm on the first three of the ten folds. C and D correspond to the first fold, E and F to the second fold, and G and H to the third fold.

**Table S1. Description of diagnostic categories in SPARK cohort.**

| Diagnostic category | Diagnostic sub-category | Variable | Definition |
|---|---|---|---|
| behave | behave | behav_adhd | ADHD (Attention Deficit-Hyperactivity Disorder) or ADD |
| behave | behave | behav_conduct | Conduct Disorder |
| behave | behave | behav_intermitt_explos | Intermittent Explosive Disorder |
| behave | behave | behav_odd | Oppositional Defiant Disorder |
| growth | growth | growth_low_wt | Difficulty gaining weight |
| growth | growth | growth_macroceph | Large head size (macrocephaly) |
| growth | growth | growth_microceph | Small head size (microcephaly) |
| growth | growth | growth_obes | Obesity |
| growth | growth | growth_short | Short stature |
| mood | mood | mood_anx | Anxiety disorder, such as panic, phobia, agoraphobia, or generalized anxiety disorder (GAD) except for social anxiety |
| mood | mood | mood_bipol | Bipolar (Manic-Depressive) Disorder |
| mood | mood | mood_dep | Depression or dysthymia |
| mood | mood | mood_dmd | Disruptive Mood Dysregulation Disorder |
| mood | mood | mood_hoard | Hoarding |
| mood | mood | mood_ocd | Obsessive-Compulsive Disorder |
| mood | mood | mood_sep_anx | Separation Anxiety |
| mood | mood | mood_soc_anx | Social Anxiety Disorder/Social Phobia |
| sleep | sleep | sleep_dx | Sleep Disorder or sleep problem diagnosed by a professional |
| birth | birth_bone | birth_def_bone | Branching Question: Birth defects of bones, hands or feet |
| birth | birth_bone | birth_def_bone_club | Clubbed foot |
| birth | birth_bone | birth_def_bone_miss | Missing or malformed bones |
| birth | birth_bone | birth_def_bone_polydact | Extra fingers and/or extra toes |
| birth | birth_bone | birth_def_bone_spine | Spine deformity |
| birth | birth_cleft | birth_def_cleft_lip | Cleft lip |
| birth | birth_cleft | birth_def_cleft_palate | Cleft palate |
| birth | birth_cns | birth_def_cns | Branching Question: Brain and spinal cord birth defects |
| birth | birth_cns | birth_def_cns_brain | Brain malformation/abnormality (shown on MRI) |
| birth | birth_cns | birth_def_cns_myelo | Spina bifida/myelomeningocele (baby born with open spine, spinal cord outside of the body) |
| birth | birth_fac | birth_def_fac | Branching Question: Facial birth defect |
| birth | birth_gi | birth_def_gastro | Branching Question: Gastrointestinal (GI) birth defects |

| | | | |
|---|---|---|---|
| birth | birth_gi | birth_def_gi_esoph_atres | Esophageal atresia (no connection between the esophagus and stomach) |
| birth | birth_gi | birth_def_gi_hirschprung | Hirschsprung disease |
| birth | birth_gi | birth_def_gi_intest_malrot | Intestinal malrotation |
| birth | birth_gi | birth_def_gi_pylor_sten | Pyloric stenosis (blockage from stomach to small intestine) |
| birth | birth_thorac | birth_def_thorac | Branching Question: Heart or lung birth defect |
| birth | birth_thorac | birth_def_thorac_cdh | Congenital diaphragmatic hernia |
| birth | birth_thorac | birth_def_thorac_heart | Congenital heart disease/defect |
| birth | birth_thorac | birth_def_thorac_lung | Lung malformation |
| birth | birth_urogen | birth_def_urogen | Branching Question: Urinary or genital birth defect |
| birth | birth_urogen | birth_def_urogen_hypospad | Hypospadias (in boys, urinary opening is in the wrong place) |
| birth | birth_urogen | birth_def_urogen_renal | Kidney malformation (for example horseshoe kidney) |
| birth | birth_urogen | birth_def_urogen_renal_agen | Missing kidney |
| birth | birth_urogen | birth_def_urogen_uter_agen | Missing uterus |
| eat | eat | eating_disorder | Eating Disorder |
| eat | eat | feeding_dx | Feeding/eating problems |
| neuro | neuro | neuro_inf | Brain infection such as bacterial meningitis, encephalitis |
| neuro | neuro | neuro_lead | Lead poisoning |
| neuro | neuro | neuro_sz | Seizure disorder or epilepsy |
| neuro | neuro | neuro_tbi | Traumatic brain injury (hospitalized) |
| visaud | visaud | visaud_blind | Blindness |
| visaud | visaud | visaud_catar | Cataract |
| visaud | visaud | visaud_deaf | Deafness/hearing loss |
| visaud | visaud | visaud_strab | Strabismus |
| development | development | dev_lang_dis | Language delay or language disorder |
| development | development | dev_ld | Learning disability (LD, learning disorder, including reading, written expression, math, or NVLD (Nonverbal learning disability)) |
| development | development | dev_motor | Motor delay (e.g., delay in walking) or developmental coordination disorder |
| development | development | dev_mutism | Mutism |
| development | development | dev_soc_prag | Social (Pragmatic) Communication Disorder |
| development | development | dev_speech | Speech articulation problems |

Data come from Basic Medical Screening dataset in SPARK phenotype dataset v9. The column Variable shows the variable name in original dataset. For birth complications, outcomes within one system were grouped as a single sub-diagnosis. For instance, if a child presented with both birth_def_cns_brain and birth_def_cns_myelo, we categorized this as a central nervous system (CNS) complication, treating it as one comorbidity within the birth category.

**Table S2. Description and statistics of quantitative measures in SPARK cohort.**

| Measure | Full name | Variable | Description | ASD-preterm (n) | ASD-term (n) | Non-ASD-preterm (n) |
|---|---|---|---|---|---|---|
| CBCL_1-5 | Child Behavior Checklist 1.5-5 years old | total_problems_t_score | Total problems T score | 309 | 1681 | 0 |
| CBCL_6-18 | Child Behavior Checklist 6-18 years old | total_problems_t_score | Total problems T score | 1101 | 7879 | 0 |
| DCDQ | Developmental Coordination Disorder Questionnaire | final_score | Final score | 4179 | 27829 | 0 |
| RBS-R | Repetitive Behavior Scale-Revised | total_final_score | Total final score | 5568 | 37653 | 0 |
| SCQ | Social-Communication Questionnaire - Lifetime | final_score | Final score | 7636 | 50527 | 2411 |
| IQ | Intelligence Quotient | fsiq_score | Overall Full Scale IQ Score | 415 | 2833 | 0 |

The column Variable shows the variable name in original dataset. The statistical numbers represent the individual counts with available data in each subgroup.

**Table S3. Description of diagnostic categories in SSC cohort.**

| Diagnostic category | Dataset | Variable | Definition |
|---|---|---|---|
| birth | medhx_fam_birth_defects | abnormal_shape_polydactyly_proband | Abnormal shape of hands, feet, arms, or legs including Polydactyly - Proband |
| birth | medhx_fam_birth_defects | cleft_lip_palate_proband | Cleft lip/palate - Proband |
| birth | medhx_fam_birth_defects | congenital_heart_defect_proband | Congenital heart defect - Proband |
| birth | medhx_fam_birth_defects | kidney_defect_proband | Kidney defect - Proband |
| birth | medhx_fam_birth_defects | open_spine_proband | Open spine - Proband |
| birth | medhx_fam_birth_defects | other_birth_defect_proband | Other birth defect - Proband |
| development | medhx_fam_language | communication_disorder_proband | Communication Disorder, NOS - Proband |
| development | medhx_fam_language | expressive_lang_disorder_proband | Expressive Language Disorder - Proband |
| development | medhx_fam_language | mixed_expressive_disorder_proband | Mixed Expressive/Receptive Language Disorder - Proband |
| development | medhx_fam_language | other_lang_disorder_proband | Other Language Disorders - Proband |
| development | medhx_fam_language | pragmatics_lang_disorder_proband | Pragmatics Language Disorder - Proband |
| development | medhx_fam_language | receptive_lang_disorder_proband | Receptive Language Disorder - Proband |
| development | medhx_fam_language | reduced_articulation_proband | Reduced articulation/pronunciation - Proband |
| development | medhx_fam_language | speech_delay_proband | Speech delay requiring therapy - Proband |
| development | medhx_fam_language | stuttering_proband | Stuttering - Proband |
| development | medhx_fam_other_developmental | math_disorder_proband | Math Disorder - Proband |
| development | medhx_fam_other_developmental | mental_retardation_proband | Mental Retardation - Proband |
| development | medhx_fam_other_developmental | nonverbal_disorder_proband | Nonverbal Learning Disorder - Proband |
| development | medhx_fam_other_developmental | other_learning_delay_proband | Other Learning delay/disorder - Proband |

| | | | |
|---|---|---|---|
| development | medhx_fam_other_developmental | reading_disorder_proband | Reading Disorder - Proband |
| development | medhx_fam_other_developmental | written_expression_disorder_proband | Disorder of Written Expression - Proband |
| neuro | medhx_fam_neurological | cerebral_palsy_proband | Cerebral Palsy - Proband |
| neuro | medhx_fam_neurological | congenital_rubella_proband | Congenital Rubella - Proband |
| neuro | medhx_fam_neurological | cranial_nerve_disorder_proband | Cranial Nerve Disorder - Proband |
| neuro | medhx_fam_neurological | hydrocephalus_proband | Hydrocephalus - Proband |
| neuro | medhx_fam_neurological | landau_kleffner_syndrome_proband | Landau Kleffner Syndrome - Proband |
| neuro | medhx_fam_neurological | migraines_proband | Migraines - Proband |
| neuro | medhx_fam_neurological | neurofibromatosis_proband | Neurofibromatosis - Proband |
| neuro | medhx_fam_neurological | seizures_proband | Seizures - Proband |
| neuro | medhx_fam_neurological | tuberous_sclerosis_proband | Tuberous Sclerosis - Proband |
| behave | medhx_fam_psychiatric1 | attention_deficit_proband | Attention Deficit Disorder/Attention Deficit Hyperactivity Disorder - Proband |
| behave | medhx_fam_psychiatric1 | behavior_disorder_proband | Behavior Disorder - Proband |
| behave | medhx_fam_psychiatric1 | other_psychotic_disorder_proband | Other Psychotic Disorders - Proband |
| behave | medhx_fam_psychiatric1 | pica_proband | Pica - Proband |
| behave | medhx_fam_psychiatric1 | tourettes_proband | Tourette's Disorder/Chronic Motor or vocal tic disorder/Tic disorder, NOS - Proband |
| mood | medhx_fam_psychiatric1 | schizophrenia_proband | Schizophrenia - Proband |
| mood | medhx_fam_psychiatric2 | adjustment_disorder_proband | Adjustment Disorder - Proband |
| mood | medhx_fam_psychiatric2 | anxiety_disorder_proband | Anxiety Disorder - Proband |
| mood | medhx_fam_psychiatric2 | bipolar_disorder_proband | Bipolar Disorder/Manic Depressive - Proband |
| mood | medhx_fam_psychiatric2 | depressive_disorder_proband | Depressive Disorder - Proband |
| mood | medhx_fam_psychiatric2 | dysthymic_disorder_proband | Dysthymic Disorder, Mood Disorder NOS - Proband |

| | | | |
|---|---|---|---|
| mood | medhx_fam_psychiatric2 | obsessive_compulsive_proband | Obsessive Compulsive Disorder - Proband |
| mood | medhx_fam_psychiatric2 | other_psych_disorder2_proband | Other Psych Disorder 2 - Proband |
| mood | medhx_fam_psychiatric2 | personality_disorder_proband | Personality Disorder - Proband |
| mood | medhx_fam_psychiatric2 | post_traumatic_stress_proband | Post Traumatic Stress Disorder - Proband |
| mood | medhx_fam_psychiatric2 | social_phobia_proband | Social Phobia - Proband |
| eat | medhx_fam_psychiatric1 | eating_disorder_proband | Eating Disorder - Proband |
| eat | medhx_perinatal | feeding_difficulty | Feeding difficulty |
| growth | medhx_perinatal | physical_anomalies_heart | Heart Defect |
| growth | medhx_perinatal | physical_anomalies_kidney | Kidney Defect |
| growth | medhx_perinatal | physical_anomalies_other | Other Defect |
| growth | medhx_perinatal | physical_anomalies_polydactyly | If yes specify: - Abnormal shape hands, feet, arms, or legs including Polydactyly |
| visaud | medhx_hearing_vision_allergies | hearing | Has a Hearing Problem |
| visaud | medhx_hearing_vision_allergies | vision | Has a Vision Problem? |

Data come from datasets listed in column Dataset in SSC phenotype dataset. The column Variable shows the variable name in original dataset.

**Table S4. Description and statistics of quantitative measures in SSC cohort.**

| Measure | Full name | Variable | Description | ASD-preterm (n) | ASD-term (n) |
|---|---|---|---|---|---|
| CBCL_2-5 | Child Behavior Checklist 1.5-5 years old | total_problems_t_score | Total Problems T Score | 44 | 394 |
| CBCL_6-18 | Child Behavior Checklist 6-18 years old | total_problems_t_score | Total Problems T Score | 111 | 1082 |
| DCDQ | Developmental Coordination Disorder Questionnaire | total | Total score | 153 | 1425 |
| SCQ | Social-Communication Questionnaire - Lifetime | summary_score | Summary_score | 136 | 1268 |
| IQ | Intelligence Quotient | fs_deviation_score | Full Scale Deviation IQ Score | 107 | 1156 |

The column Variable shows the variable name in original dataset. The statistical numbers represent the individual counts with available data in each subgroup.

**Table S5. Quality control and parameters used in polygenic risk score calculation.**

| **Parameter used in Plink for QC** | | |
|---|---|---|
| **Parameter** | **Threshold** | **Description** |
| --mind | 0,04 | Genotyping threshold of individuals (less than 3 standard deviations of mean missing call rate) |
| --geno | 0,01 | Missingness of variants |
| --hwe | 1,00E-06 | Hardy–Weinberg equilibrium of variants |
| --maf | 0,01 | Minor allele frequency threshold of variants |
| --indep-pairwise | 200 50 0.25 | Linkage disequlibrium of variants |
| --het | | Individuals with heterozygosity rates out of the range of ±3 standard deviations of mean heterozygosity rate are removed |
| **Dafult Parameter used in PRScs** | | |
| **Parameter** | **Threshold** | **Description** |
| --a | 1 | PARAM_A |
| --b | 0,5 | PARAM_B |
| --phi | 0,01 | PARAM_PHI |
| --n_iter | 1000 | MCMC_ITERATIONS |
| --n_burnin | 500 | MCMC_BURNIN |
| --thin | 5 | MCMC_THINNING_FACTOR |

**Table S6. Feature selected in machine learning model construction.**

| **Feature** | **Description (data type)** |
|---|---|
| Sex | Sex assigned at birth (female\|male) |
| Birth complication Exist | Labeled as 1 if individual has any one of the birth complications (1\|0) |
| Gestational at age | Gestational age at birth (number) |
| Oxygen Deprivation at Birth | Labeled as 1 if individual had insufficient oxygen at birth with NICU stay (1\|0) |
| Polygenic Risk Score (ASD) | Standarized ASD Polygenic Risk Score (number) |
| Recessive NDD-InherVar Number | The number of inherited variants on exonic recessive neurodevelopmental disorder (NDD) -related genes (number) |
| Dominant NDD-InherVar Number | The number of inherited variants on exonic dominant NDD genes (number) |
| Total DNV Number | The number of de novo varinanats (DNV) (number) |
| NDD-DNV Number | The number of DNV on NDD genes (number) |
| Autosomes-Exonic-DNV Number | The number of DNV on autosomes and only considering protein-coding genes (number) |
| Sum CADD Score (NDD-DNV) | The sum of the Combined Annotation Dependent Depletion (CADD) scores of DNV on NDD genes (number) |
| Avg CADD Score (NDD-DNV) | The average of CADD scores of DNV on NDD genes (number) |
| LOF Number | The number of LOF DNV (number) |

* Variants mentioned in this table were identified in genome sequencing.

**Table S7. The values of hyperparameter tuning for machine learning models.**

| Algorithm | Parameter | Value |
|---|---|---|
| XGBoost | nrounds | 100, 200, 300, 400 |
| | eta | 0.03, 0.05, 0.1, 0.2, 0.3 |
| | max_depth | 3, 6, 9, 12 |
| | gamma | 0, 1, 3, 5 |
| | colsample_bytree | 0.3, 0.5, 0.75 |
| | min_child_weight | 1, 3 |
| | subsample | 0,8 |
| | scale_pos_weight | 0,6 |
| Random Forest | mtry | 1, 3, 5, 7, 10, 15, 20 |
| | ntree | 1000 |
| | nodesize | 1 |
| Linear SVM | C | 0, 0.01, 0.05, 0.1, 0.25, 0.5, 0.75, 1, 1.25, 1.5, 1.75, 2 |
| | preProcess | "center","scale" |

**Table S8. The prevalence and odds ratio of diagnostic categories compared between ASD-preterm and ASD-term in the SPARK cohort.**

| Diagnostic category | Phenotype | Diagnosed number | Prevalence | OR (95% CI) | FDR-p | GEE: OR (95% CI) | GEE: FDR-p |
|---|---|---|---|---|---|---|---|
| Behavior | Preterm | 3900 | 0.42 | 1.19 (1.14–1.25) | 5.04E-15 | 1.27 (1.21–1.33) | 3.16E-24 |
| | Term | 24809 | 0.38 | | | | |
| Birth | Preterm | 529 | 0.06 | 2.18 (1.97–2.4) | 5.61E-55 | 2.25 (2.03–2.49) | 1.20E-54 |
| | Term | 1774 | 0.03 | | | | |
| Development | Preterm | 6597 | 0.72 | 1.5 (1.43–1.57) | 4.88E-61 | 2.45 (2.27–2.63) | 2.37E-43 |
| | Term | 40896 | 0.63 | | | | |
| Eat | Preterm | 2070 | 0.23 | 1.71 (1.62–1.8) | 4.12E-86 | 1.67 (1.58–1.77) | 2.43E-75 |
| | Term | 9456 | 0.15 | | | | |
| Growth | Preterm | 1748 | 0.19 | 2.18 (2.06–2.31) | 9.24E-158 | 2.23 (2.1–2.37) | 1.37E-150 |
| | Term | 6310 | 0.10 | | | | |
| Mood | Preterm | 3245 | 0.35 | 1.1 (1.05–1.15) | 4.39E-05 | 1.31 (1.25–1.38) | 7.80E-27 |
| | Term | 21548 | 0.33 | | | | |
| Neuro | Preterm | 923 | 0.10 | 1.64 (1.52–1.76) | 1.61E-38 | 1.75 (1.62–1.89) | 4.41E-46 |
| | Term | 4150 | 0.06 | | | | |
| Sleep | Preterm | 2417 | 0.26 | 1.45 (1.38–1.52) | 3.44E-47 | 1.49 (1.42–1.57) | 2.25E-51 |
| | Term | 12856 | 0.20 | | | | |
| Visaud | Preterm | 944 | 0.10 | 2.02 (1.87–2.17) | 3.88E-76 | 2.15 (1.99–2.33) | 5.50E-84 |
| | Term | 3493 | 0.05 | | | | |

FDR-p values are adjusted p-values by False Discovery Rate method. ORs with 95% CI are given among ASD individuals born preterm vs term. Both unadjusted odds ratios and those adjusted for covariates (sex and age) and family clustering using GEE models are provided. Diagnostic categories are described in table S1.

**Table S9. Statistical test of differences of prevalence between different preterm stages (For SPARK).**

| Diagnosis | Comparison | p.Chisq | p.adj.Chisq (FDR adjustmenmt) | p.adj.tau (FDR adjustmenmt) |
|---|---|---|---|---|
| Behave | Extremely preterm : Very preterm | 0,404 | 0,577 | 0,3 |
| Behave | Extremely preterm : Moderate preterm | 0,817 | 0,817 | |
| Behave | Extremely preterm : Late preterm | 0,651 | 0,723 | |
| Behave | Extremely preterm : Term | 0,0154 | 0,0385 | |
| Behave | Very preterm : Moderate preterm | 0,473 | 0,591 | |
| Behave | Very preterm : Late preterm | 0,0842 | 0,168 | |
| Behave | Very preterm : Term | 2,83E-05 | 0,000142 | |
| Behave | Moderate preterm : Late preterm | 0,349 | 0,577 | |
| Behave | Moderate preterm : Term | 0,000264 | 0,00088 | |
| Behave | Late preterm : Term | 1,07E-07 | 1,07E-06 | |
| Growth | Extremely preterm : Very preterm | 0,0518 | 0,0576 | 0,038 |
| Growth | Extremely preterm : Moderate preterm | 0,0104 | 0,0149 | |
| Growth | Extremely preterm : Late preterm | 3,33E-07 | 6,66E-07 | |
| Growth | Extremely preterm : Term | 9,01E-48 | 4,51E-47 | |
| Growth | Very preterm : Moderate preterm | 0,577 | 0,577 | |
| Growth | Very preterm : Late preterm | 0,00343 | 0,00572 | |
| Growth | Very preterm : Term | 2,91E-38 | 7,28E-38 | |
| Growth | Moderate preterm : Late preterm | 0,0141 | 0,0176 | |
| Growth | Moderate preterm : Term | 1,79E-41 | 5,97E-41 | |
| Growth | Late preterm : Term | 9,31E-89 | 9,31E-88 | |
| Mood | Extremely preterm : Very preterm | 0,0298 | 0,093 | 0,92 |
| Mood | Extremely preterm : Moderate preterm | 0,497 | 0,621 | |
| Mood | Extremely preterm : Late preterm | 0,366 | 0,523 | |
| Mood | Extremely preterm : Term | 0,914 | 0,914 | |

| | | | | |
|---|---|---|---|---|
| Mood | Very preterm : Moderate preterm | 0,0737 | 0,147 | |
| Mood | Very preterm : Late preterm | 0,0372 | 0,093 | |
| Mood | Very preterm : Term | 0,000387 | 0,00387 | |
| Mood | Moderate preterm : Late preterm | 0,888 | 0,914 | |
| Mood | Moderate preterm : Term | 0,199 | 0,332 | |
| Mood | Late preterm : Term | 0,0033 | 0,0165 | |
| Sleep | Extremely preterm : Very preterm | 0,372 | 0,413 | 1 |
| Sleep | Extremely preterm : Moderate preterm | 0,0544 | 0,109 | |
| Sleep | Extremely preterm : Late preterm | 0,271 | 0,339 | |
| Sleep | Extremely preterm : Term | 0,0027 | 0,00675 | |
| Sleep | Very preterm : Moderate preterm | 0,268 | 0,339 | |
| Sleep | Very preterm : Late preterm | 0,991 | 0,991 | |
| Sleep | Very preterm : Term | 4,30E-07 | 1,43E-06 | |
| Sleep | Moderate preterm : Late preterm | 0,13 | 0,217 | |
| Sleep | Moderate preterm : Term | 3,54E-14 | 1,77E-13 | |
| Sleep | Late preterm : Term | 2,64E-31 | 2,64E-30 | |
| Birth | Extremely preterm : Very preterm | 0,425 | 0,531 | 0,3 |
| Birth | Extremely preterm : Moderate preterm | 0,733 | 0,733 | |
| Birth | Extremely preterm : Late preterm | 0,208 | 0,297 | |
| Birth | Extremely preterm : Term | 6,66E-10 | 1,66E-09 | |
| Birth | Very preterm : Moderate preterm | 0,577 | 0,641 | |
| Birth | Very preterm : Late preterm | 0,00678 | 0,0136 | |
| Birth | Very preterm : Term | 3,46E-20 | 1,73E-19 | |
| Birth | Moderate preterm : Late preterm | 0,0301 | 0,0502 | |
| Birth | Moderate preterm : Term | 8,51E-20 | 2,84E-19 | |
| Birth | Late preterm : Term | 1,18E-30 | 1,18E-29 | |
| Eat | Extremely preterm : Very preterm | 0,00608 | 0,00869 | 0,038 |
| Eat | Extremely preterm : Moderate preterm | 0,000225 | 0,000375 | |

| | | | | |
|---|---|---|---|---|
| Eat | Extremely preterm : Late preterm | 2,69E-08 | 5,38E-08 | |
| Eat | Extremely preterm : Term | 2,27E-33 | 1,14E-32 | |
| Eat | Very preterm : Moderate preterm | 0,386 | 0,386 | |
| Eat | Very preterm : Late preterm | 0,022 | 0,0275 | |
| Eat | Very preterm : Term | 3,14E-19 | 1,05E-18 | |
| Eat | Moderate preterm : Late preterm | 0,183 | 0,203 | |
| Eat | Moderate preterm : Term | 8,31E-18 | 2,08E-17 | |
| Eat | Late preterm : Term | 4,45E-44 | 4,45E-43 | |
| Neuro | Extremely preterm : Very preterm | 0,0213 | 0,0237 | 0,038 |
| Neuro | Extremely preterm : Moderate preterm | 0,00549 | 0,00686 | |
| Neuro | Extremely preterm : Late preterm | 2,44E-09 | 4,88E-09 | |
| Neuro | Extremely preterm : Term | 3,05E-26 | 3,05E-25 | |
| Neuro | Very preterm : Moderate preterm | 0,713 | 0,713 | |
| Neuro | Very preterm : Late preterm | 0,00162 | 0,0027 | |
| Neuro | Very preterm : Term | 3,55E-14 | 8,88E-14 | |
| Neuro | Moderate preterm : Late preterm | 0,0032 | 0,00457 | |
| Neuro | Moderate preterm : Term | 4,16E-15 | 1,39E-14 | |
| Neuro | Late preterm : Term | 8,27E-16 | 4,14E-15 | |
| Visaud | Extremely preterm : Very preterm | 7,46E-07 | 1,07E-06 | 0,038 |
| Visaud | Extremely preterm : Moderate preterm | 3,30E-09 | 5,50E-09 | |
| Visaud | Extremely preterm : Late preterm | 1,64E-25 | 5,47E-25 | |
| Visaud | Extremely preterm : Term | 1,26E-77 | 1,26E-76 | |
| Visaud | Very preterm : Moderate preterm | 0,493 | 0,493 | |
| Visaud | Very preterm : Late preterm | 0,000268 | 0,000335 | |
| Visaud | Very preterm : Term | 6,68E-23 | 1,67E-22 | |
| Visaud | Moderate preterm : Late preterm | 0,00256 | 0,00284 | |
| Visaud | Moderate preterm : Term | 3,35E-22 | 6,70E-22 | |
| Visaud | Late preterm : Term | 4,49E-28 | 2,24E-27 | |

| | | | | |
|---|---|---|---|---|
| Development | Extremely preterm : Very preterm | 0,00168 | 0,0028 | 0,15 |
| Development | Extremely preterm : Moderate preterm | 0,00915 | 0,0131 | |
| Development | Extremely preterm : Late preterm | 1,66E-05 | 3,32E-05 | |
| Development | Extremely preterm : Term | 7,82E-18 | 3,91E-17 | |
| Development | Very preterm : Moderate preterm | 0,441 | 0,49 | |
| Development | Very preterm : Late preterm | 0,514 | 0,514 | |
| Development | Very preterm : Term | 5,27E-09 | 1,32E-08 | |
| Development | Moderate preterm : Late preterm | 0,0772 | 0,0965 | |
| Development | Moderate preterm : Term | 1,50E-14 | 5,00E-14 | |
| Development | Late preterm : Term | 2,87E-33 | 2,87E-32 | |

Kendall's rank tau test was performed with ranking from extremely preterm to term order, and p-values were adjusted by FDR method.

**Table S10. The prevalence and odds ratio of multimorbidity in diagnostic categories compared between ASD-preterm and ASD-term in the SPARK cohort.**

| Multimorbudity | Phenotype | Disagnosed number | Prevalence | OR (95% CI) | FDR-p | GEE: OR (95% CI) | GEE: FDR-p |
|---|---|---|---|---|---|---|---|
| 0 | Preterm | 766 | 0.08 | 0.60 (0.55–0.65) | 3.32E-39 | 0.57 (0.53–0.62) | 4.02E-43 |
| | Term | 8585 | 0.13 | | | | |
| 1 | Preterm | 2438 | 0.27 | 0.73 (0.70–0.77) | 8.22E-35 | 0.70 (0.67–0.74) | 3.14E-43 |
| | Term | 21421 | 0.33 | | | | |
| 2 | Preterm | 2076 | 0.23 | 0.93 (0.88–0.98) | 0.004 | 0.95 (0.90–0.997) | 0.039 |
| | Term | 15559 | 0.24 | | | | |
| 3 | Preterm | 1695 | 0.18 | 1.19 (1.13–1.26) | 1.10E-09 | 1.24 (1.17–1.31) | 2.48E-13 |
| | Term | 10349 | 0.16 | | | | |
| 4 | Preterm | 1084 | 0.12 | 1.48 (1.38–1.59) | 3.61E-29 | 1.55 (1.44–1.66) | 1.08E-33 |
| | Term | 5369 | 0.08 | | | | |
| >=5 | Preterm | 1137 | 0.12 | 2.31 (2.16–2.48) | 3.59E-126 | 2.45 (2.27–2.63) | 8.96E-127 |
| | Term | 3738 | 0.06 | | | | |

FDR-p values are adjusted p-values by False Discovery Rate method. ORs with 95% CI are given among ASD individuals born preterm vs term when focusing on each multimorbidity number. Both unadjusted odds ratios and those adjusted for covariates (sex and age) and family clustering using GEE models are provided. Diagnostic categories are described in table S1.

**Table S11. FDR-adjusted p values in Post-hoc comparison of Chi-square test for the burden of multimorbidities in different groups (for SPARK).**

| **Number of multimorbidities** | **0** | **1** | **2** | **3** | **4** | **>=5** |
|---|---|---|---|---|---|---|
| Whole preterm vs Term | 6,63E-39 | 2,47E-34 | 0,026 | 5,48E-09 | 1,44E-28 | 3,59E-126 |
| Extremely preterm vs Very preterm | 0,46 | 1 | 1 | 1 | 1 | 0,33 |
| Extremely preterm vs Moderate preterm | 0,014 | 1 | 1 | 0,30 | 1 | 0,36 |
| Extremely preterm vs Late preterm | 0,002 | 0,01 | 1 | 0,098 | 1 | 7,99E-06 |
| Extremely preterm vs Term | 2,33E-10 | 5,92E-09 | 0,64 | 0,0003 | 0,0002 | 3,27E-40 |
| Very preterm vs Moderate preterm | 0,92 | 1 | 1 | 0,64 | 1 | 1 |
| Very preterm vs Late preterm | 0,33 | 0,13 | 1 | 0,22 | 1 | 0,060 |
| Very preterm vs Term | 1,85E-08 | 2,70E-08 | 0,37 | 0,0003 | 2,16E-06 | 6,41E-29 |
| Moderate preterm vs Late preterm | 1 | 0,21 | 1 | 1 | 1 | 0,012 |
| Moderate preterm vs Term | 2,91E-06 | 3,63E-09 | 0,63 | 0,28 | 6,96E-07 | 1,14E-38 |
| Late preterm vs Term | 3,08E-22 | 1,78E-15 | 0,25 | 0,0007 | 2,52E-18 | 1,14E-63 |

The multimorbidity counts the number of diagnostic categories described in Table S1.

**Table S12. The prevalence and odds ratio of diagnostic categories compared between ASD-preterm and non-ASD-preterm in the SPARK cohort.**

| Diagnostic category | Phenotype | Diagnosed number | Prevalence | OR (95% CI) | FDR-p | GEE: OR (95% CI) | GEE: FDR-p |
|---|---|---|---|---|---|---|---|
| Behavior | ASD | 3900 | 0.42 | 3.03 (2.73–3.36) | 1.04E-103 | 2.43 (2.19–2.71) | 3.12E-103 |
| | non-ASD | 529 | 0.20 | | | | |
| Birth | ASD | 529 | 0.06 | 1.93 (1.53–2.45) | 2.71E-08 | 1.78 (1.4–2.27) | 2.71E-08 |
| | non-ASD | 83 | 0.03 | | | | |
| Development | ASD | 6597 | 0.72 | 8.77 (7.94–9.71) | 0 | 9.1 (8.19–10.1) | 0 |
| | non-ASD | 607 | 0.22 | | | | |
| Eat | ASD | 2070 | 0.23 | 5.57 (4.67–6.7) | 6.73E-95 | 5.88 (4.91–7.05) | 1.51E-94 |
| | non-ASD | 134 | 0.05 | | | | |
| Growth | ASD | 1748 | 0.19 | 2.02 (1.77–2.32) | 1.00E-25 | 2.02 (1.76–2.32) | 1.29E-25 |
| | non-ASD | 281 | 0.10 | | | | |
| Mood | ASD | 3245 | 0.35 | 3 (2.68–3.36) | 1.18E-86 | 2.65 (2.36–2.99) | 2.12E-86 |
| | non-ASD | 416 | 0.15 | | | | |
| Neuro | ASD | 923 | 0.10 | 4.13 (3.26–5.33) | 1.64E-34 | 3.9 (3.03–5.02) | 2.45E-34 |
| | non-ASD | 71 | 0.03 | | | | |
| Sleep | ASD | 2417 | 0.26 | 6.94 (5.82–8.36) | 8.40E-126 | 6.87 (5.72–8.24) | 3.78E-125 |
| | non-ASD | 132 | 0.05 | | | | |
| Visaud | ASD | 944 | 0.10 | 2.23 (1.85–2.7) | 8.68E-18 | 2.25 (1.85–2.73) | 9.77E-18 |
| | non-ASD | 132 | 0.05 | | | | |

FDR-p values are adjusted p-values by False Discovery Rate method. ORs with 95% CI are given among ASD vs non-ASD individuals born preterm. Both unadjusted odds ratios and those adjusted for covariates (sex and age) and family clustering using GEE models are provided. Diagnostic categories are described in table S1. Totally 9196 ASD and 2706 non-ASD.

**Table S13. The prevalence and odds ratio of multimorbidity in diagnostic categories compared between ASD-preterm and non-ASD-preterm in the SPARK cohort.**

| Multimorbudity | Phenotype | Diagnosed number | Prevalence | OR (95% CI) | FDR-p | GEE: OR (95% CI) | GEE: FDR-p |
|---|---|---|---|---|---|---|---|
| 0 | ASD | 766 | 0.08 | 0.09 (0.08–0.1) | 0 | 0.1 (0.09–0.11) | 0 |
| | Non-ASD | 1357 | 0.50 | | | | |
| 1 | ASD | 2438 | 0.27 | 0.92 (0.84–1.02) | 0.097 | 0.98 (0.88–1.08) | 0.097 |
| | Non-ASD | 761 | 0.28 | | | | |
| 2 | ASD | 2076 | 0.23 | 2.04 (1.81–2.31) | 2.31E-30 | 1.92 (1.69–2.18) | 2.31E-30 |
| | Non-ASD | 338 | 0.12 | | | | |
| 3 | ASD | 1695 | 0.18 | 4.51 (3.76–5.45) | 6.45E-67 | 4.16 (3.45–5.01) | 6.45E-67 |
| | Non-ASD | 129 | 0.05 | | | | |
| 4 | ASD | 1084 | 0.12 | 5.17 (4.07–6.7) | 1.76E-46 | 4.89 (3.8–6.3) | 1.76E-46 |
| | Non-ASD | 68 | 0.03 | | | | |
| >=5 | ASD | 1137 | 0.12 | 7.04 (5.38–9.42) | 2.37E-56 | 6.59 (4.97–8.74) | 2.37E-56 |
| | Non-ASD | 53 | 0.02 | | | | |

FDR-p values are adjusted p-values by False Discovery Rate method. ORs with 95% CI are given among ASD vs non-ASD individuals born preterm when focusing on each multimorbidity number. Both unadjusted odds ratios and those adjusted for covariates (sex and age) and family clustering using GEE models are provided. Diagnostic categories are described in table S1. Totally 9196 ASD and 2706 non-ASD. Totally 157 preterm and 1479 term.

**Table S14. The prevalence and odds ratio of diagnostic categories compared between ASD-preterm and ASD-term in the SSC cohort**

| Diagnostic category | Phenotype | Diagnosed number | Prevalence | OR (95% CI) | FDR-p | GEE: OR (95% CI) | GEE: FDR-p |
|---|---|---|---|---|---|---|---|
| Behavior | Preterm | 32 | 0.20 | 1.45 (0.95–2.18) | 0.21 | 1.54 (1.01–2.35) | 0.12 |
| | Term | 222 | 0.15 | | | | |
| Birth | Preterm | 5 | 0.03 | 0.8 (0.27–1.84) | 0.68 | 0.78 (0.31–1.98) | 0.75 |
| | Term | 60 | 0.04 | | | | |
| Development | Preterm | 79 | 0.50 | 0.94 (0.68–1.31) | 0.73 | 0.94 (0.67–1.3) | 0.75 |
| | Term | 766 | 0.52 | | | | |
| Eat | Preterm | 53 | 0.34 | 2.58 (1.79–3.68) | 7.7E-07 | 2.55 (1.78–3.66) | 2.85E-06 |
| | Term | 244 | 0.16 | | | | |
| Growth | Preterm | 14 | 0.09 | 2.02 (1.06–3.58) | 0.08 | 1.98 (1.08–3.62) | 0.10 |
| | Term | 69 | 0.05 | | | | |
| Mood | Preterm | 7 | 0.04 | 0.8 (0.33–1.65) | 0.68 | 0.88 (0.39–1.96) | 0.75 |
| | Term | 83 | 0.06 | | | | |
| Neuro | Preterm | 8 | 0.05 | 0.75 (0.33–1.47) | 0.68 | 0.76 (0.36–1.59) | 0.75 |
| | Term | 101 | 0.07 | | | | |
| Visaud | Preterm | 35 | 0.22 | 1.15 (0.77–1.7) | 0.68 | 1.27 (0.84–1.92) | 0.53 |
| | Term | 295 | 0.20 | | | | |

FDR-p values are adjusted p-values by False Discovery Rate method. ORs with 95% CI are given among ASD individuals born preterm vs term. Both unadjusted odds ratios and those adjusted for covariates (sex and age) and family clustering using GEE models are provided. Diagnostic categories are described in table S1. Totally 157 preterm and 1479 term.

**Table S15. The prevalence and odds ratio of multimorbidity in diagnostic categories compared between ASD-preterm and ASD-term in the SSC cohort.**

| Multimorbudity | Phenotype | Disagnosed number | Prevalence | OR (95% CI) | FDR-p | GEE: OR (95% CI) | GEE: FDR-p |
|---|---|---|---|---|---|---|---|
| 0 | Preterm | 31 | 0.20 | 0.8 (0.52–1.19) | 0.34 | 0.78 (0.51–1.18) | 0.30 |
| | Term | 350 | 0.24 | | | | |
| 1 | Preterm | 57 | 0.36 | 0.77 (0.54–1.08) | 0.21 | 0.75 (0.53–1.05) | 0.16 |
| | Term | 631 | 0.43 | | | | |
| 2 | Preterm | 41 | 0.26 | 1.18 (0.8–1.7) | 0.40 | 1.22 (0.83–1.78) | 0.31 |
| | Term | 342 | 0.23 | | | | |
| 3 | Preterm | 19 | 0.12 | 1.71 (0.99–2.8) | 0.18 | 1.76 (1.04–2.97) | 0.14 |
| | Term | 111 | 0.08 | | | | |
| >=4 | Preterm | 9 | 0.06 | 1.96 (0.88–3.92) | 0.18 | 2.05 (0.98–4.29) | 0.14 |
| | Term | 45 | 0.03 | | | | |

FDR-p values are adjusted p-values by False Discovery Rate method. ORs with 95% CI are given among ASD individuals born preterm vs term when focusing on each multimorbidity number. Both unadjusted odds ratios and those adjusted for covariates (sex and age) and family clustering using GEE models are provided. Diagnostic categories are described in table S1. Totally 157 preterm and 1479 term.

**Table S16. P-values from GEE models for the correlation between multimorbidity and DNV count (SPARK cohort).**

| | GS_DNV | GS_LOF | GS_DNV_NDD | GS_LOF_NDD | ES_DNV | ES_LOF | ES_DNV_NDD | ES_LOF_NDD |
|---|---|---|---|---|---|---|---|---|
| **Model 1: multimorbidity ~ Variant_count + sex, data = Whole** | | | | | | | | |
| Variant_count | 0.16 (-) | 0.037 (+) | 0,2 | 5.3e-06 (+) | 1.1e-05 (+) | 1.9e-09 (+) | 6.6e-14 (+) | <2e-16 (+) |
| **Model 2: Preterm ~ Variant_count * multimorbidity + sex, data = ASD** | | | | | | | | |
| Variant_count | 0,6 | 0,904 | 0,217 | 0,71 | 0,68 | 0,98 | 0,36 | 0,34 |
| multimorbidity | 0,91 | 5.5e-07 (+) | 0,54 | 4.9e-07 (+) | 3.9e-07 (+) | 9.2e-14 (+) | 5.6e-12 (+) | 1e-14 (+) |
| Variant_count:multimorbidity | 0,22 | 0,904 | 0.040 (+) | 0,61 | 0,98 | 0,97 | 0,84 | 0,66 |
| **Model 3: ASD ~ Variant_count * multimorbidity + sex, data = Preterm** | | | | | | | | |
| Variant_count | 0,053 | 0,59 | 0.0066 (-) | 0,596 | 0.049 (+) | 0,59 | 0,58 | 0,48 |
| multimorbidity | 0,77 | 9.8e-12 (+) | 0,33 | 1.5e-12 (+) | 8.6e-08 (+) | 2.2e-12 (+) | 1.1e-10 (+) | 1.1e-13 (+) |
| Variant_count:multimorbidity | 0,22 | 0,18 | 0.0325 (+) | 0,424 | 0,47 | 0,26 | 0,85 | 0,92 |

The +/- after p value reflects the positive/negative estimation of variables. GS = genome sequencing, ES = exome sequencing, NDD = NDD-related genes.

**Table S17. Performance metrics of machine learning model used to predict ASD diagnosis in all individuals.**

| Algorithm | Accuracy | 95% CI | AUC | Sensitivity | Specificity | F1-score |
|---|---|---|---|---|---|---|
| XGBoost | 0.686 | (0.672, 0.699) | 0.677 | 0.791 | 0.519 | 0.755 |
| Random forest | 0.675 | (0.661, 0.688) | 0.656 | 0.821 | 0.443 | 0.756 |
| SVM | 0.695 | (0.679, 0.710) | 0.650 | 0.802 | 0.522 | 0.764 |
